# Leveraging DNA Methylation to Predict Treatment Response in Major Depressive Disorder: A Critical Review

**DOI:** 10.1101/2023.12.01.23299308

**Authors:** Jan Dahrendorff, Glenn Currier, Monica Uddin

**Author notes:** Corresponding author: Address correspondence to: Monica Uddin, PhD University of South Florida, 3720 Spectrum Blvd., Suite 304.

## Abstract

**Background:** Major depressive disorder (MDD) is a debilitating and prevalent mental disorder with a high disease burden. Despite a wide array of different treatment options, many patients do not respond to initial treatment attempts. Selection of the most appropriate treatment remains a significant clinical challenge in psychiatry, highlighting the need for the development of biomarkers with predictive utility. Recently, the epigenetic modification DNA methylation (DNAm) has emerged to be of great interest as a potential predictor of MDD treatment outcomes. Here we review efforts to date that seek to identify DNAm signatures associated with treatment response in individuals with MDD.

**Methods:** Searches were conducted in the databases PubMed, Scopus, and Web of Science with the concepts and keywords major depressive disorder, DNA methylation, antidepressants, psychotherapy, cognitive behavior therapy, electroconvulsive therapy, transcranial magnetic stimulation, and brain stimulation therapies.

**Results:** We identified 31 studies implicating DNAm patterns associated with MDD treatment outcomes. The majority of studies (*N*=25) are focused on selected target genes exploring treatment outcomes in pharmacological treatments (*N=22*) with a few studies assessing treatment response to electroconvulsive therapy (*N=3*). Additionally, there are few genome-scale efforts (*N=6*) to characterize DNAm patterns associated with treatment outcomes.

**Limitations:** There is a relative dearth of studies investigating DNAm patterns in relation to psychotherapy, electroconvulsive therapy, or transcranial magnetic stimulation; importantly, most existing studies have limited sample sizes.

**Conclusions:** Given the heterogeneity in both methods and results of studies to date, there is a need for additional studies before existing findings can inform clinical decisions.

## 1. Introduction

Major depressive disorder (MDD) is a debilitating and highly prevalent mental disorder characterized by a range of symptoms including depressed mood, loss of interest in activities that once brought enjoyment, impairment of cognitive functioning, fatigue, sleep disturbances, and psychomotor changes (Association, 2013). MDD is the most prevalent mental health disorder in the United States with current estimates of the past year and lifetime prevalence of around 10% and 20%, respectively (Goodwin et al., 2022; Hasin et al., 2018). Globally, the World Health Organization estimates a lifetime prevalence of 4.4% making MDD also the most common mental disorder (World Health Organization, 2017). MDD is highly recurrent (Hardeveld et al., 2013; Moffitt et al., 2010) and associated with a substantial disease burden (Jia, Zack, Thompson, Crosby, & Gottesman, 2015). Many people suffering from the disorder can face a reduction in quality-adjusted life expectancy of around 30 years; as such, MDD is among the diseases with the highest loss of quality-adjusted life expectancy (Jia et al., 2015).

Additionally, individuals suffering from MDD are at an increased risk to develop a range of comorbidities (Arnaud et al., 2022) including cardiovascular diseases (Brunner et al., 2014; Surtees et al., 2008), metabolic disorders (Rotella & Mannucci, 2013; Vancampfort et al., 2015), neurodegenerative diseases (Diniz, Butters, Albert, Dew, & Reynolds, 2013; Ownby, Crocco, Acevedo, John, & Loewenstein, 2006), and autoimmune disorders(Andersson et al., 2015).

Although there have been significant efforts to illuminate the molecular underpinnings of MDD, the etiology of MDD remains poorly understood. Despite the role of genetic factors in the etiology of MDD (reviewed in (Flint, 2023; Shadrina, Bondarenko, & Slominsky, 2018)) the identified heritability estimates--around 40% (Kendler, Gatz, Gardner, & Pedersen, 2006; Kendler, Ohlsson, Lichtenstein, Sundquist, & Sundquist, 2018) ––are lower in comparison to other psychiatric disorders (e.g. schizophrenia 81%, Sullivan et al., 2003; bipolar disorder 85% (McGuffin et al., 2003) and ADHD 82% (Chang, Lichtenstein, Asherson, & Larsson, 2013))suggesting that additional factors beyond DNA sequence might mediate MDD pathogenesis. Recent work has proposed that epigenetic mechanisms play a role in mediating the risk of MDD after environmental exposures (Klengel & Binder, 2015). Broadly, epigenetic alterations influence gene expression and translation via chemical modifications that regulate either chromatin structure or modify the accessibility of DNA, resulting in altered transcriptional activity of the locus (J. K. Kim, Samaranayake, & Pradhan, 2009). These processes include DNA methylation (DNAm), histone modifications, and small non-coding RNAs, among others. Among known epigenetic mechanisms, DNAm is perhaps the most studied epigenetic modification. A large body of literature documents a range of environmental factors that can lead to changes in DNAm patterns including exposure to pollutants and toxins (Hou, Zhang, Wang, & Baccarelli, 2012), smoking (Breitling, Yang, Korn, Burwinkel, & Brenner, 2011) aging (Bjornsson et al., 2008; Wong et al., 2010), diet (summarized here (Maugeri & Barchitta, 2020)), medication (Kringel, Malkusch, & Lotsch, 2021; Lotsch et al., 2013) physical activity (Sailani et al., 2019), early life experiences (Cecil, Zhang, & Nolte, 2020; Klengel et al., 2013; Martins et al., 2021; Weaver, Meaney, & Szyf, 2006) and stress exposure(van der Knaap et al., 2014). Similarly, aberrant DNAm can modulate the transcriptional activity of genes, enabling variation in gene expression patterns that can potentially mediate how an individual responds to environmental stimuli (Provencal et al., 2020) or even contribute to the pathogenesis of diseases (Nishiyama & Nakanishi, 2021; van den Oord et al., 2022).

DNAm has emerged to be of interest as a predictive molecular biomarker in MDD treatment response (Gasso et al., 2017; Neyazi et al., 2018; Wang, Zhang, et al., 2018). In such pharmaco-epigenetic studies, DNAm is measured in patients before, and in some efforts, also post-completion of the treatment. In parallel, repeated depression symptom measures are obtained throughout the treatment. The assessment of DNAm at baseline and after completion of the treatment can thereby be utilized to characterize epigenomic variation associated with treatment outcomes and in turn, inform the development of tools with predictive utility (Domschke et al., 2014; Ju et al., 2019; L. Li et al., 2021; Takeuchi et al., 2017). Advancing the development of such predictive tools could potentially guide future treatment decisions and ultimately shift trial-and-error treatment approaches toward more optimal treatment choices.

Several pharmacological and non-pharmacological treatment options for MDD are currently available including pharmacotherapy, psychotherapy, and brain stimulation treatments (Reviewed in (Gartlehner et al., 2017; Meerman, Ter Hark, Janzing, & Coenen, 2022)), and often a combination of such treatments is utilized to maximize treatment outcomes. Despite the variety of existing MDD therapeutics, a large proportion of patients do not respond to initial treatment attempts (Trivedi et al., 2006; Wang, Zhang, et al., 2018), with treatments leading to remission in only approximately one-third of the patients (Trivedi et al., 2006; Undurraga & Baldessarini, 2012; Warden, Rush, Trivedi, Fava, & Wisniewski, 2007). Similarly, between a third and 55% of patients with MDD are classified as suffering from treatment-resistant depression, a particularly severe form of depression in which two or more treatment attempts with antidepressant medication have previously been unsuccessful (Souery et al., 2007; Thomas et al., 2013; Trivedi et al., 2006). Identification of the right treatment for a patient is largely based on trying out different treatments, which usually takes a long time to achieve a clinical response in the patient, often times causing substantial hardship to those seeking relief from their symptoms (Garcia-Gonzalez et al., 2017; Papakostas, 2008) (Figure 1 for Illustration). The ability to provide treatments that are most likely to elicit a positive response in any individual patient remains an elusive goal in psychiatry that could perhaps be informed by findings from DNAm studies. To advance this goal, here, we review and summarize existing studies investigating DNAm-based epigenetic signatures associated with treatment outcomes in patients with MDD. Given that the majority of epigenetic studies in relation to treatment outcomes are examining DNAm, we will focus our review on this particular epigenetic modification.

**Figure 1.**
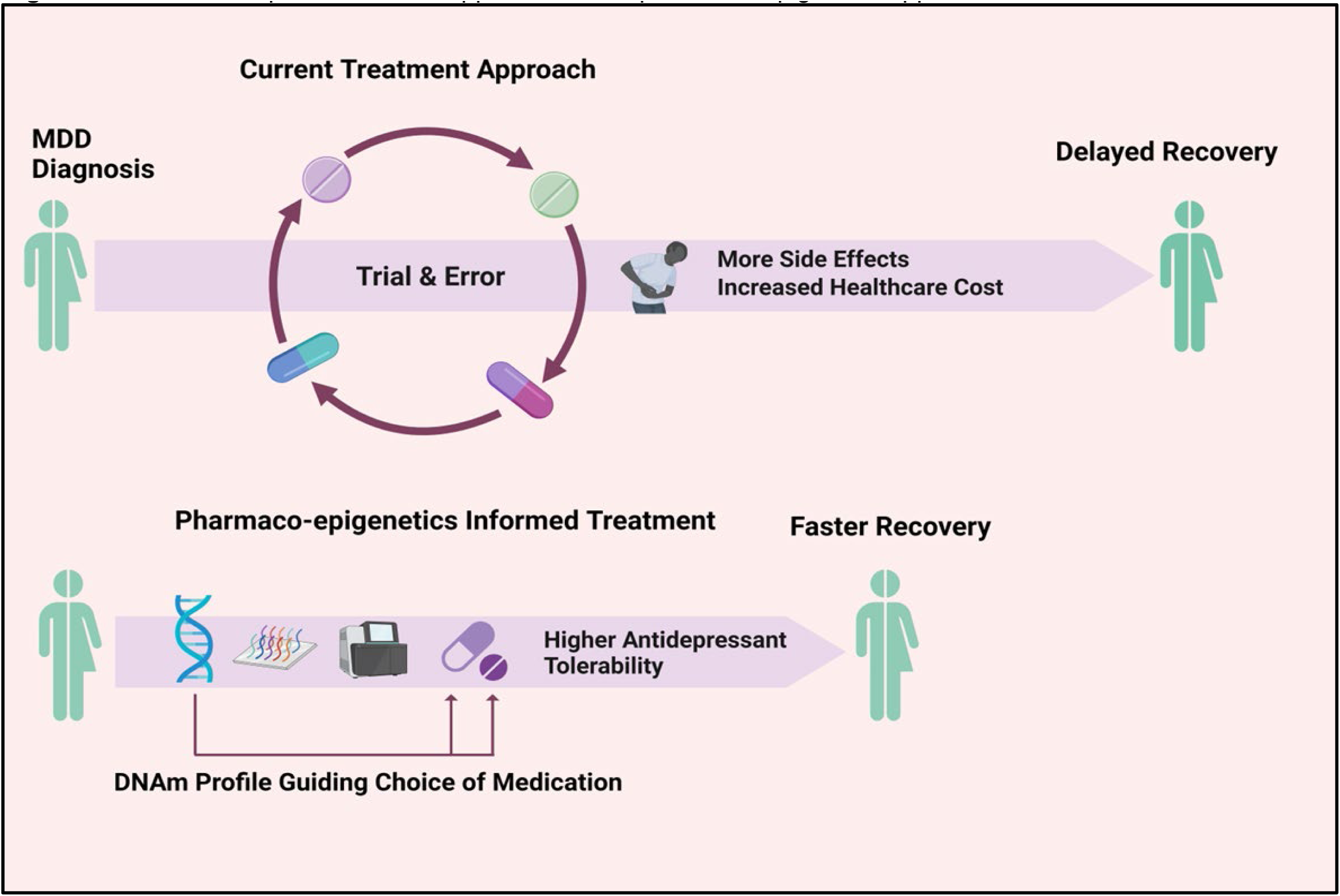
Schematic representation of opportunities for pharmaco-epigenetic approaches in MDD treatment.

## 2. Materials and Method

The searches for this review were conducted in the databases PubMed (United States National Library of Medicine at the National Institute of health), Scopus (Elsevier), and Web of Science (Clarivate) to identify primary research articles on DNAm associated with treatment response in MDD. The following search terms for this review included: Major depressive disorder, DNA methylation, antidepressants, psychotherapy, cognitive behavior therapy, brain stimulation therapy, electroconvulsive therapy (ECT), and transcranial magnetic stimulation (TMS). The search included primary research articles in English. Studies focusing on epigenetic mechanisms other than DNAm, conference abstracts, review papers, murine and cell culture studies investigating DNAm associated with treatment responses were excluded from the search. The searches in PubMed were conducted on September 7^th^, 2023, while the Scopus and Web of Science searches were conducted on September 7^th^, 2023.

The PubMed search identified 52 articles; Scopus yielded 48 articles while Web of Science returned 49 results. Additionally, two records that were identified in the reference list of a previous review paper were considered significant and subsequently added to the search results. In total, 151 articles were identified and imported into EndNote reference management software (Clarivate). After the removal of duplicated articles, 54 articles remained for initial review. Next, review articles, book chapters, case reports, case studies, and murine and cell culture studies were removed after screening the titles. Additionally, studies focused on characterizing DNAm alterations induced by MDD treatments that have not assessed treatment outcomes, and studies focused on other psychiatric disorders were excluded from our results. This filtering procedure removed 21 sources, producing a total of 33 primary research articles that underwent a full-text review. The full-text review led to the exclusion of two additional articles: One literature review and one article not evaluating treatment outcomes, leaving 31 research articles for inclusion in the study (Figure 2 for illustration of literature search).

**Figure 2.**
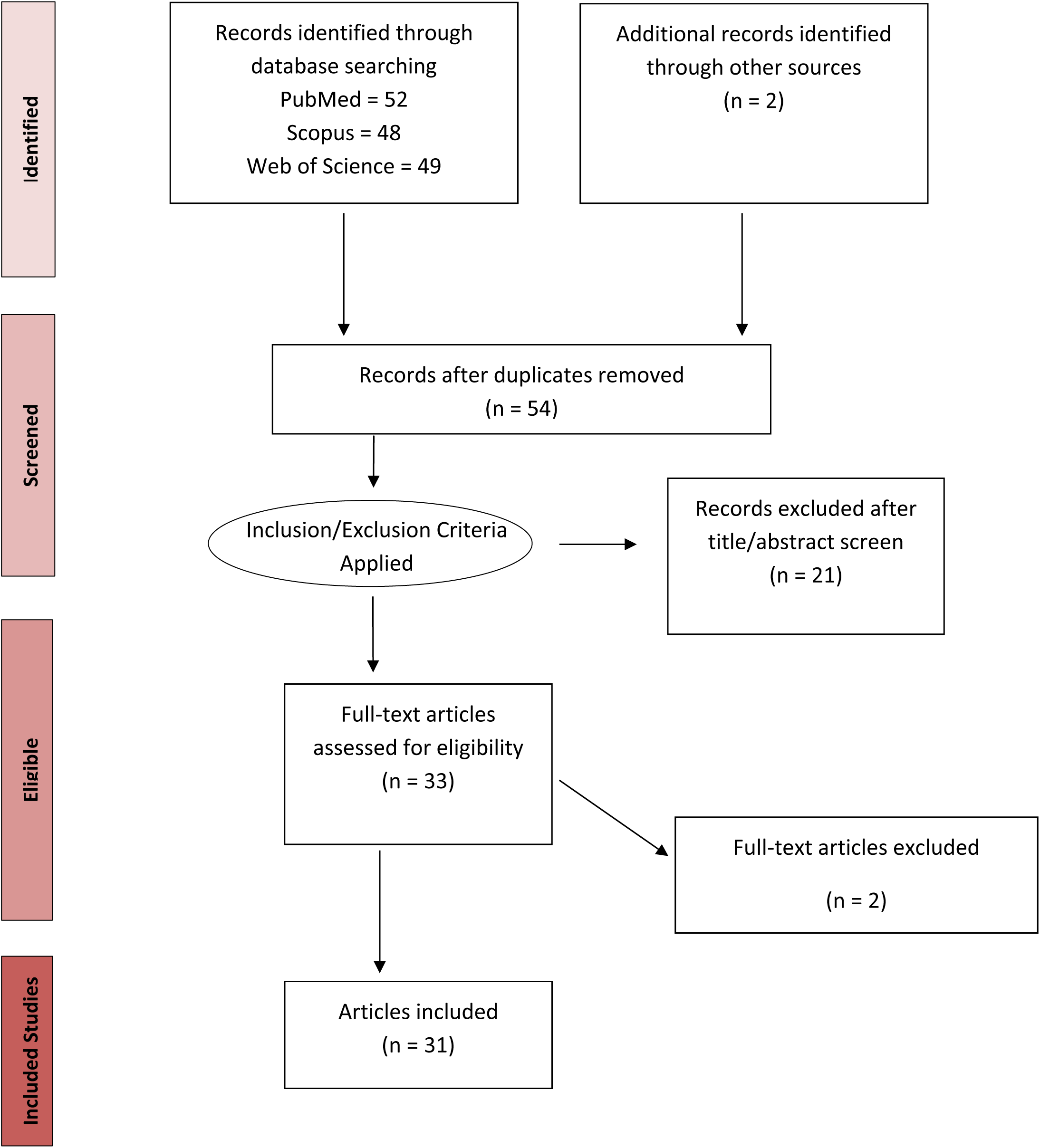
Literature search diagram showing the filtering and article selection process

## 3. Results

Figure 3 presents the results of the literature search process employed in this study. Table 1 presents a summary of all studies included in this review. To date, most epigenetic investigations (*N* = 25) of MDD treatment responses are focused on selected target genes that have been well characterized in the context of MDD including solute carrier family 6 member 4 (*SLC6A4)*, brain-derived neurotrophic factor (*BDNF)*, interleukin (*IL*)*-1*, *IL-6* & *IL-11,* monoamine oxidase *A (MAOA),* 5-hydroxytryptamine receptor *(HTR1A/1B)* and tryptophan hydroxylase 2 *(TPH2)*. However, studies focusing on genome-scale analyses of methylation are also emerging and have become more frequent in the last few years (*N=6*). The majority of such genome-scale efforts have investigated pharmacological treatments of MDD as the treatment modality (*N=4*)(Engelmann et al., 2022; Ju et al., 2019; Martinez-Pinteno et al., 2021; Takeuchi et al., 2017) with only a few focused on ECT (*N=2)(Moschny, Zindler, et al., 2020; Sirignano et al., 2021)*.

**Figure 3.**
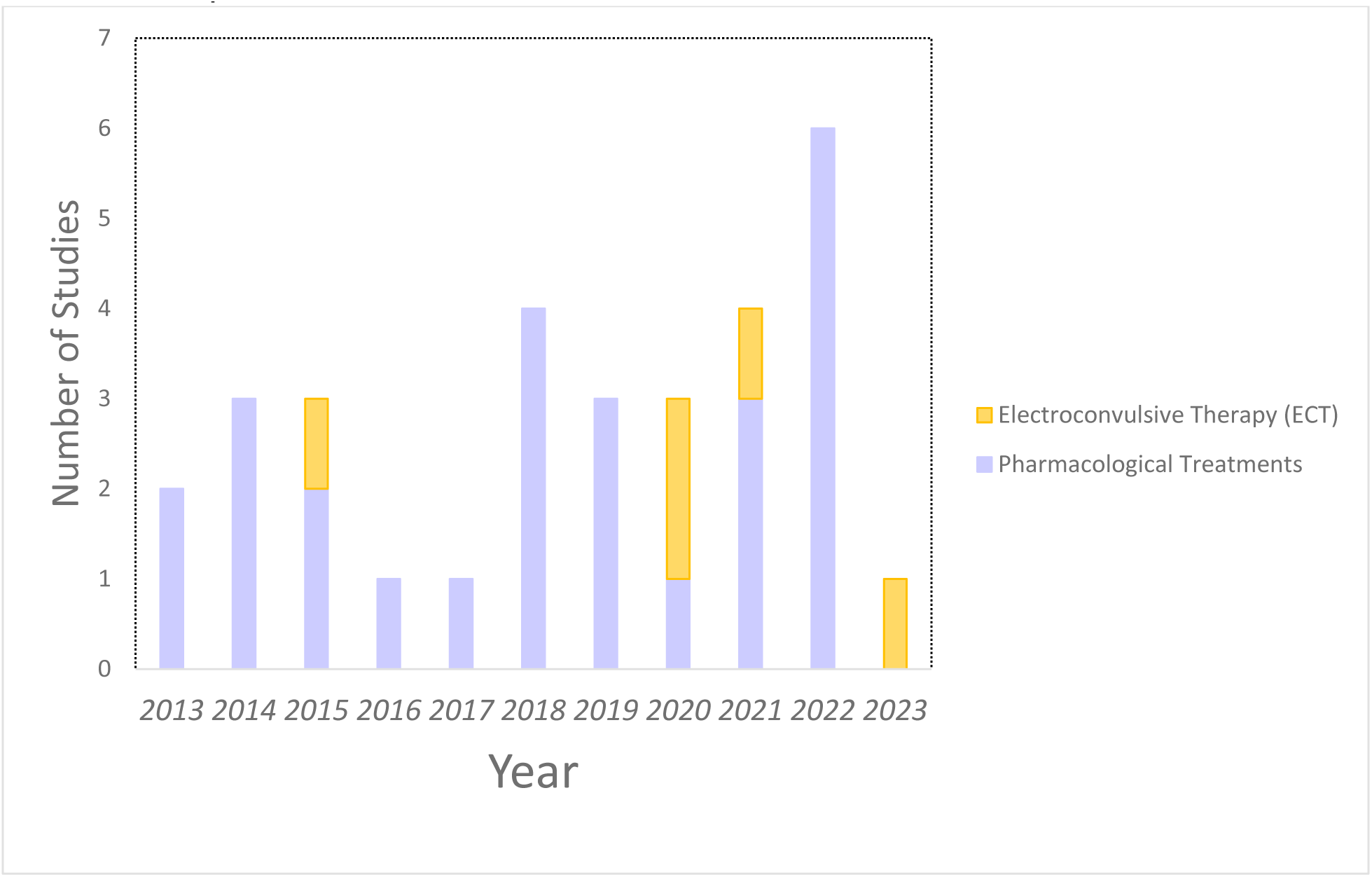
Increase of studies investigating DNAm signatures in MDD associated with a treatment response over the last decade

**Table 1.** Overview of studies characterizing DNA methylation profiles associated with treatment response in Major depressive disorder.

| Author & Date | Sample | Antidepressant Treatment | Treatment Duration and Outcome Measure | Genomic Loci Assessed | Tissue Examined | Method to assess Methylation | Direction of Effect in Relation to Treatment Response |
| --- | --- | --- | --- | --- | --- | --- | --- |
| Domschke et al. (2014) | 94 MDD (females n=61, males n=33), European ancestry | Various antidepressants/Escitalopram only subsample (n= 61) | 6 weeks, HAM-D% change | 9 CpGs of *SLC6A4* exon 1A | Peripheral Blood | Targeted direct bisulfite sequencing (measured at baseline) | ↓meth average ↓response mainly conferred by CpG 2 (Bonferroni corrected) |
| Domschke et al. (2015) | 94 MDD (females n=61, males n=33), European ancestry | Various antidepressants/ Escitalopram - only subsample (n=61) | 6 weeks, HAM-D% change | 43 CpGs in *MAO-A* | Peripheral Blood | Targeted direct bisulfite sequencing (measured at baseline) | trend for ↓meth in two CpGs ↓response in females (nominally significant) |
| Dragonov et al. (2019) | 155 MDD (females n=105, males n=48), European ancestry | Various antidepressants | 4 weeks, Response = MSM ≤7 | 5 CpG sites of *IL6*, 6 sites of *IL1-β* and 2 of *IL6R* | Peripheral Blood | Bisulfite-pyrosequencing (measured at baseline) | ↑meth *IL6R* CpG ↑response (nominally significant) |
| Engelmann et al. (2022) | 80 MDD (female n=46, males n=34), European ancestry | Escitalopram, venlafaxine, lithium | 4 weeks, HAM-D% change | EWAS/ Genome-scale | Whole blood | Illumina EPIC BeadChip (measured at baseline) | No epigenome-scale significant differentially methylated positions, 18 ↑meth DMRs, 2 ↓meth DMRs; strongest in *SORBS2*, *CYP2C18* ↑response (Šidák corrected) |
| Gao et al. (2022) | 98 MDD (female n=55, male n=43), Asian ancestry | Various antidepressants | 2 weeks, HAM-D% change | 181 CpGs sites of *HTR1A* and *HTR1B*. | Peripheral Blood | Bisulfite Amplicon Sequencing (measured at baseline) | ↓meth *HTR1A* CpG 208 & *HTR1B* CpG 65 ↓response (nominally significant) |
| Gasso et al. (2017) | 57 MDD, (no sex breakdown of participants in MDD cohort) adolescents between 10-17 years, European ancestry | Fluoxetine, comedication with antipsychotics & (or) benzodiazepines or mood stabilizers | 12 weeks, CDI change | 7 CpG sites in *HTR1B* promoter | Peripheral Blood | Bisulfite-pyrosequencing (measured at baseline) | ↓meth *HTR1B* promoter ↑response (nominally significant) |
| Hsieh et al. (2019) | 51 MDD, 62 controls (female n=25 male n=25 in case and controls), Asian ancestry | Various antidepressants & benzodiazepines | 4 weeks, HAM-D% change | 14 CpG sites in *BDNF* exon IX | Peripheral Blood | Bisulfite-pyrosequencing (measured at baseline) | ↑meth *BDNF* CpG 24 & CpG 324 ↑response (nominally significant) |
| Iga et al. (2016) | 28 MDD, (female n=20, male n=8), Asian ancestry | Various antidepressants | 8 weeks, HAM-D% change | 9 CpG sites in *SLC6A4* promoter | Peripheral Blood | Bisulfite-pyrosequencing (measured at baseline & completion of treatment) | trend for ↓meth *SLC6A4* CpG 2 ↑response (Bonferroni corrected) |
| Ju et al. (2019) | 177 MDD (female n=110 male, n=67), Majority of European ancestry | Escitalopram | 8 weeks, MADRS% change | EWAS/ Genome-scale | Peripheral Blood | Infinium Methylation EPIC Beadchip (measured at baseline) | ↓meth two DMP in *CHN2* and one in DMP in *JAK2* ↑response, one DMP in *CHN2* replicated in an external cohort |
| Kang et al. (2013) | 108 MDD (female n=75 male, n=33), Asian ancestry | Various antidepressants | 12 weeks, HAM-D% change | 7 CpGs in *SLC6A4* promoter | Peripheral Blood | Bisulfite-pyrosequencing (measured at baseline) | trend for ↑meth CpG 2 ↓response (nominally significant) |
| Kleimann et al. (2015) | 11 patients (female n=5, male n=6), European ancestry | ECT | 3.5 weeks, MADRS% change, BDI-II change | *BDNF* promoter I, IV, and VI CpGs | Peripheral Blood | Bisulfite-pyrosequencing (measured at baseline, throughout & after treatment) | ↓meth *BDNF* exon I promoter ↑response (nominally significant) |
| Kim et al. (2015) | 127 MDD & acute coronary syndrome (no sex breakdown of participants in MDD) | Escitalopram | 24-weeks, HAM-D% change | 9 CpGs sites in *BDNF* exon VI | Peripheral Blood leukocytes | Bisulfite-pyrosequencing (measured at baseline & completion of treatment) | ↑meth one *BDNF* CpG & average ↓response |
|  | cohort), Asian ancestry |  |  |  |  |  |  |
| Li et al. (2021) | 291 MDD (female n=189, male n=102), Asian ancestry | Various antidepressants ( SSRIs, SNRIs, NaSSAs, SARIs) | 6 weeks, HAM-D% change | 194 CpG sites spanning six CpG Islands of *BDNF* | Peripheral Blood | Targeted NGS-based methylation analysis (measured at baseline) | ↑meth *BDNF* CpG 140<br>↑response (FDR corrected) |
| Lieb et al. (2018) | 561 MDD (female n=315, male n=246) [199 Severe MDD HAM-17 scores ≥ 25 (female n=118, male n=81), European ancestry | Escitalopram for 2 weeks, venlafaxine in non-responders, lithium added after 28 days in non-responders | 8 weeks, HAM-D% change | *BDNF* Exon IV promoter and *p11* promoter | Peripheral Blood | Targeted direct bisulfite sequencing (measured at baseline) | ↑meth *BDNF* CpG-87 in patients with severe MDD<br>↑response (uncorrected for multiple testing) |
| Maier et al. (2023) | 31 MDD (female n=20, male n=11), European ancestry | ECT | 3-4 weeks, BDI-II and MADRS% change | 43 CpG Sites in *NR3C1* and 28 CpG Sites in *POMC* | PBMCs | Sanger sequencing of bisulfite-converted DNA<br>Bisulfite-pyrosequencing (measured at baseline, throughout & after treatment) | ↓meth *NR3C1*<br>↑response (nominally significant) |
| Martinez-Pinteño et al. (2021) | 22 MDD (female n=19, male n=3), children and adolescents between 13-17 years, European ancestry | Fluoxetine | 8 weeks, CDI%- & CGI-I% change | EWAS/ Genome-scale | Peripheral Blood | Illumina Infinium Methylation EPIC BeadChip (measured at baseline) | ↑meth CpG sites *RHOJ* &<br>↓meth *OR2L13*<br>↓response (FDR corrected) |
| Moon et al. (2022) | 221 MDD (female n=173, male n=48), Asian ancestry | Various SSRIs (Escitalopram, Fluoxetine, Paroxetine, Sertraline | 6 weeks, HAM-D% change | Nine CpG sites in the promoter region of *SLC6A4* | Peripheral blood | Bisulfite-pyrosequencing (measured at baseline & completion of treatment) | No DMPs identified<br>↑meth CpG4, ↓meth CpG3 & CpG5 observed after treatment completion (nominally significant) |
| Moschny et al. (2020)a | 87 MDD (female n=45, male n=42), European ancestry | ECT | HAM-D% Change, MADRS% change | 39 CpG sites in *t-PA* and 35 CpGs in *PAI-1* | PBMCs | Sanger sequencing of bisulfite-converted DNA (measured at baseline, throughout & after treatment) | No differences in baseline *t-PA* or *PAI-1* meth (Šidák corrected) |
| Moschny et al. (2020)b | 17 MDD (female n=10, male n=7), European ancestry | ECT | 4 weeks, BDI-II and MADRS% change | EWAS/ Genome-scale | PBMCs | Illumina NextSeq 550 (measured at baseline & completion of treatment) | ↓meth *RNF175*, ↑meth *RNF213*, ↓meth *TBC1D14*, ↑meth *TMC5*, ↑meth *WSCD1*, ↓meth *AC018685.2*, ↓meth *AC098617.1*, ↑meth *CLCN3P1* ↑ response |
| Okada et al. (2014) | 50 MDD 50 controls (female n=25, male n=25 in case and controls), Asian Ancestry | Paroxetine, fluvoxamine, milnacipran | 6 weeks, HAM-D% change | 81 CpGs in *SLC6A4* promoter | Peripheral Blood | Targeted methylation array (measured at baseline & completion of treatment) | ↑meth *SLC6A4* CpG 3 ↑response (Bonferroni corrected) |
| Powell et al. (2013) | 113 MDD (female n=67, male n=46), European ancestry | 80 Escitalopram, 33 Nortriptyline | 12 weeks, MADRS% change | CpG island of *interleukin-11* loci. | Peripheral Blood | Targeted methylation assessment (measured at baseline) | ↓meth CpG 5 ↑ response ↑meth CpG4 ↑response ↑meth CpG 11 x rs1126757 GG ↑response (FDR corrected) |
| Sirignano et al. (2021) | 34 MDD (female n=16, male n=18), European ancestry | ECT | HAM-D% change | EWAS/ Genome-scale & 799 candidate gene CpGs | Peripheral Blood | Illumina Infinium Methylation EPIC array (measured at baseline & completion of treatment) | ↑meth *FKBP5* CpG at follow-up ↑response in continuous response model (FDR corrected) No differences in baseline meth after correction for multiple comparisons) |
| Schiele et al. (2021) | 236 MDD (female n=138, male n= 98) European ancestry | Various anti-depressants | 6 weeks, HAM-D% change | 9 CpG sites located in the *SLC6A4* promoter region | Blood cells | Direct sequencing of sodium bisulfite-treated DNA (measured at baseline) | ↓meth CpG ↓response, similar trend in longitudinal replication cohort (n=110) (nominally significant) |
| Shen et al. (2020) | 291 MDD (female n=187, male n=104) 1000 healthy controls, | Various anti-depressants | HAM-D% Change, 2 weeks | 38 CpG sites of *TPH2* | Peripheral blood | NGS-based multiple Targeted CpG methylation analysis method | ↑meth 2 CpGs in male & ↑meth 1 CpG in female ↓response (uncorrected for multiple testing) ↑meth 2 CpGs x ↑ early life stress ↓response (FDR corrected) |
|  | Asian ancestry |  |  |  |  | (measured at baseline) |  |
| Tadic et al. (2014) | 39 MDD, (female n=22, male n=17) European ancestry | Fluoxetine, Venflaxine, TCAs | 6 weeks, HAM-D% change | 12 CpG sites of *BDNF* exon IV promoter | PBMCs | Direct sequencing of sodium bisulfite-treated DNA | ↓meth CpG 87 at baseline<br>↓response |
| Takeuchi et al. (2018) | 20 MDD (female n=11, male n=9), Asian ancestry | Paroxetine | 6 weeks, HAM-D% change | EWAS/ Genome-scale | Peripheral Blood | Infinium HumanMethylation450 BeadChip (measured at baseline) | ↑meth *PPFIA4* CpG cg00594917<br>↑meth *HS3ST1* CPG cg07260927<br>↓response (FDR corrected) |
| Wang et al. (2018)a | 85 MDD (female n=56, male n=29), Asian ancestry | Escitalopram & low dose of benzodiazepines | 8 weeks, HAM-D% change | 96 CpG sites in *HTR1A* & *HTR1B* promoter | Peripheral blood | Bisulfite Amplicon Sequencing (measured at baseline & completion of treatment) | ↓meth *HTR1A1* CpG 668 & *HTR1B1* CpG 1401<br>↓response<br>↑meth *HTR1A1* CpG 668 x Life Events Scale score<br>↓response (FDR corrected) |
| Wang et al. (2018)b | 85 MDD (female n=56, male n=29), 44 longitudinal, Asian ancestry | Escitalopram & low dose of benzodiazepines | 8 weeks, HAM-D% change | 90 CpG sites in *BDNF* | Peripheral blood | Bisulfite Amplicon Sequencing (measured at baseline & completion of treatment) | ↑meth *BDNF* at four amplicons in *BDNF*<br>↑response<br>↓average meth *BDNF*<br>↓response<br>↓Stressful life events x<br>↑meth ↑response (FDR corrected) |
| Wang et al. (2022) | 291 MDD patients (female n=189, male n=102) & 100 controls (female n=64, male n=36), Asian ancestry | Various Antidepressants (SSRI-group & non-SSRI-group) | 2 weeks, HAM-D% change | 74 CpG sites of *P11* | Peripheral blood | Bisulfite Amplicon Sequencing (measured at baseline) | ↑meth *P11-3-185* x early life stress<br>↓response (FDR corrected) |
| Xu et al. (2022)a | 291 MDD patients (female n=189, male n=102) & 100 controls (female n=64, male n=36), Asian ancestry | Various Antidepressants (SSRI-group & non-SSRI-group) | 2 weeks, HAM-D% change | 181 CpGs sites of *HTR1A* and *HTR1B*. | Peripheral blood | Bisulfite Amplicon Sequencing (measured at baseline) | ↓meth *HTR1A-2* CpG 143 + low CTQ score<br>↑response<br>↑meth *HTR1B* CpG 3-61 x *rs6298* AA/AG genotype<br>↑response |
| Xu et al. (2022)b | 87 MDD (female n=72, male n=15) & 53 controls (female n=39 male<br>n=14) Asian<br>ancestry | Various Antidepressants | 6 weeks, BDI-II ≤ 10 | 3 CpGs of *RPS6KA5* | Peripheral blood | Bisulfite-pyrosequencing (measured at baseline) | ↑average meth<br>↑response (Benjamini-Hochberg adjusted) |

### 3.1 Candidate gene DNAm studies investigating antidepressant response

The vast majority of candidate gene studies investigating DNAm patterns in association with treatment outcomes are focused on pharmacological treatments for MDD. While some studies assess epigenetic marks associated with a response to selective serotonin reuptake inhibitors (SRRIs) (*N*=9), most studies include patients undergoing a more heterogenous treatment regime utilizing a combination of tricyclic antidepressants (TCAs), selective norepinephrine reuptake inhibitors (SNRIs), or noradrenergic and specific serotonergic antidepressants (NaSSA), in addition to SRRIs. Those pharmaco-epigenetic candidate gene studies can grant insights into DNAm patterns of loci previously implicated in MDD and potentially be of predictive utility by characterizing DNAm patterns in those who respond or do not respond to a given treatment.

#### 3.1.A Monoamine-transporters, oxidases, receptors & related genes

Among the first proposed hypotheses regarding the etiology of MDD was the ‘monoamine theory’ in which serotonin (5-HT), norepinephrine, and dopamine deficiencies and imbalances were thought to underly the development of MDD (Coppen, 1967; Hamon & Blier, 2013; Schildkraut, 1965). Early findings from studies using monoamine oxidase inhibitors and TCAs potentiating 5-HT activity and in turn, ameliorating depressive symptoms, have informed decades of research investigating monoamine systems in MDD (reviewed in (Z. Li, Ruan, Chen, & Fang, 2021; Morilak & Frazer, 2004)). As a result, a vast body of literature has investigated monoamine neurotransmitter genes in MDD (reviewed in (Flint & Kendler, 2014; Shadrina et al., 2018).

The majority of such studies have analyzed genes in the serotonin pathway, e.g., the serotonin transporter *(SLC6A4)* or serotonin receptor (*HTR1A/1B*). Many antidepressant drugs exert their antidepressant effects by selectively inhibiting the function of *SLC6A4* which is to mediate the reuptake of serotonin, a key neurotransmitter in the brain. In addition, both serotonin receptor subtypes *HTR1A* and *HTR1B* play a critical role in mediating antidepressant activity and have therefore been of great research interest (Fakhoury, 2015). The chronic administration of SSRIs desensitizes the presynaptic *HTR1A* and *HTR1B* autoreceptors, decreasing the negative feedback mediated by the receptors, and in turn, stimulating activation of post synaptic serotonin receptors eliciting the antidepressant response (Chilmonczyk, Bojarski, Pilc, & Sylte, 2015; Tiger, Varnas, Okubo, & Lundberg, 2018).

Concerning pharmaco-epigenetic studies investigating candidate genes, *SLC6A4* has been the most frequently assessed locus with six studies having investigated differential DNAm associated with treatment outcomes following antidepressant treatment to date. All of the studies have examined CpGs in the promoter region of *SLC6A4.* A number of early studies was conducted in populations of Asian ancestry. The first study by Kang and colleagues (Kang et al., 2013) interrogated the methylation status of seven CpGs within the promoter of *SLC6A4* in the peripheral blood of 108 Korean patients, but only observed nominally significant differential methylation (hypermethylation) of one CpG in treatment non-responders at baseline. A study by Okada et al. (2014) conducted in Japan examined *SLC6A4* promoter methylation in the peripheral blood of patients undergoing MDD treatment and interrogated 81 CpGs in the *SLC6A4* gene using a mass spectrometry approach. The authors identified hypermethylation of CpG 3 of *SLC6A4* in treatment responders. A similar effort in Japan (Iga et al., 2016) assessed *SLC6A4* promoter methylation in 28 MDD patients undergoing treatment with various antidepressants including SSRIs and TCAs for eight weeks. Akin to the earlier findings by Kang and colleagues (Kang et al., 2013), the authors identified the differential methylation of one CpG at baseline to be nominally associated with treatment outcomes, however, Iga and colleagues describe their observations in relation to symptom improvement, with hypomethylation of one CpG associated with improvement in symptom severity following eight weeks of treatment.

In contrast, Domschke and colleagues (2014) observed reduced *SLC6A4* methylation across nine CpGs at baseline associated with an impaired response to escitalopram treatment after six weeks in 94 patients of European descent suffering from MDD. Interestingly, the seven previously investigated CpGs by Kang et al., showed the opposite direction of effect in this study: while Domschke et al. reported hypomethylation to be associated with an inhibited treatment response in their European cohort (Domschke et al., 2014), Kang and colleagues observed a relative increase in methylation—at least at one CpG site--to be associated with an impaired improvement (Kang et al., 2013). Despite these contradictory findings, the findings by Domschke and colleagues have since been supported in a recent observational study by Schiele et al. (2021), similarly investigating DNAm of nine CpG sites in the promoter region of *SLC6A4.* The authors evaluated DNAm for its association with antidepressant treatment response in the peripheral blood of 236 participants with MDD receiving various antidepressants including SSRIs, SNRIs, TCAs and NaSSA over the six-week-long in-patient treatment. Their analysis of nine CpG sites by sequencing of bisulfite-converted DNA revealed that hypomethylation of several *SLC6A4* CpGs was associated with an impaired antidepressant response, as in the earlier work by Domschke and colleagues. Their findings were replicated in another subset of patients that continued treatments with antidepressants (n= 110) (Schiele et al., 2021).

Overall, findings from studies investigating *SLC6A4* have been inconclusive with often contrasting results. The results of two studies (Iga et al., 2016; Kang et al., 2013) converge on findings of a negative correlation between promoter methylation and treatment outcomes, with Kang and colleagues reporting a trend of increased *SLC6A4* promoter CpG methylation associated with impaired treatment response while Iga and colleagues reporting lower methylation associated with symptom improvement. Conversely, two studies (Domschke et al., 2014; Schiele et al., 2021) observed *SLC6A4* hypomethylation in individuals unresponsive to antidepressant treatment while Okada and colleagues found increased *SLC6A4* promoter methylation in treatment responders. Lastly, a study in Korea (Moon et al., 2022) investigated *SLC6A4* promoter DNAm pre– and post-6 weeks of SSRI treatment but did not identify any Differentially methylated positions (DMPs) associated with treatment response, further obscuring the predictive utility of *SLC6A4* promoter methylation in relation to treatment outcomes.

Other genes of interest for pharmaco-epigenic studies in the serotonin pathway include the *Serotonin receptor subtypes (HTR1A/1B).* Four studies to date have investigated serotonin receptor subtype methylation level in association with MDD treatment response. Akin to contradictions among results from studies of the serotonin transporter gene, there have been inconsistencies among the findings in studies of *HTR1A/1B.* For example, a study by Gassó and colleagues (2017), investigated the effects of DNAm and genetic variation of the 5-hydroxytryptamine receptor 1B *(HTR1B)* gene in relation to treatment response to the SSRI fluoxetine (Gasso et al., 2017). In their study, 83 adolescent participants were evaluated for MDD (n=57), obsessive compulsive disorder (n=16) and generalized anxiety disorder (n=12) symptoms 12 weeks into their fluoxetine treatment. The authors focused on the effect of SNPs in transcription factor binding sites, proteins highly involved with the regulation of transcription and gene expression in the *HTR1B* locus, and clinical outcomes among the patients. Their analysis found the two SNPs *s9361233* and *rs9361235*, to be significantly associated with a positive treatment outcome after fluoxetine treatments. Similarly, their DNAm analysis of 7 selected CpGs in the promoter region of *HTR1B* was assessed via pyrosequencing and showed a negative correlation between positive clinical outcomes after fluoxetine treatment and average *HTR1B* promoter region methylation levels at baseline. However, the co-medication of some participants with antipsychotics, mood stabilizers, and benzodiazepines limits the interpretability of the results.

In contrast, Wang and colleagues (Wang, Lv, et al., 2018) focused on patients exclusively treated with escitalopram and observed hypomethylation at one CpG of *HTR1A* & and one CpG of *HTR1B* to be associated with an impaired respond to escitalopram. Additionally, the authors observed the interaction of recent stressful life events and hypomethylation at four CpG sites of *HTR1A/1B* to be associated with an impaired treatment response.

Another example is a recent study by Gao and colleagues (2022), who applied neuroimaging and sequencing techniques to investigate brain activity and *HTR1A/1B* DNAm with antidepressant treatment response in 300 patients to develop a model capable of predicting treatment outcomes based on their data. Their analysis examined over 116 different regions of the brain and 181 CpG sites located in the promoter region of *HTR1A* and *HTR1B* while taking 11 clinical characteristics into account. Their logistic regression model resulted in 78.57% prediction accuracy with a 0.834 area under the ROC curve (AUC). Of note, the methylation sites assessed in the study were in proximity to the CpGs investigated in the earlier study by Wang *et al* (2018a (Wang, Lv, et al., 2018)) and showed similar lower methylation levels in the group resistant to treatment, despite using a wide range of different antidepressants (SSRIs, SNRIs, and NaSSAs). Finally, Xu and colleagues (Xu et al., 2022) focused their analysis on the impact of DNAm of *HTR1A* and *HTR1B* in combination with stressful life events and childhood adversity on antidepressant treatment response. Stratifying their analysis into a SSRI and non-SSRI group, the authors assessed methylation in 181 CpG sites of the *HTR1A* and *HTR1B* genes. Hypomethylation of one *HTR1A* CpG was associated with an improved antidepressant response in patients of the non-SSRI group (treated with SNRIs and NaSSAs). Additionally, the authors observed an interaction of *HTR1A* methylation at one CpG and childhood adversity, such that hyper methylation together with high levels of childhood aversity, were significantly associated with poor treatment outcomes in both groups. Further, participants with the rs6298 SNP who also had high methylation levels of *HTR1B CpG3-61,* showed significantly better efficacy of SSRI treatment.

To date, studies of *HTR1A/1B CpG* methylation in relation to treatment outcomes are scarce, report contradicting directions of effect, and show heterogeneity in study designs, making comparisons among findings challenging. However, it is noteworthy that interaction of CpG methylation and genotype, or environmental effects such as stressful life events or childhood aversity, could jointly inform treatment outcomes as opposed to just by methylation alone.

An important regulator of monoamine levels is monoamine oxidases (MAOs) and differential methylation of this locus has been assessed in the context of antidepressant treatment response (Domschke et al., 2015). MAOs are enzymes located on the outer membranes of mitochondria responsible for the metabolism of monoamine neurotransmitters via oxidative deamination (Yeung, Georgieva, Atanasov, & Tzvetkov, 2019). For instance, serotonin is degraded by MAO-A (Tong et al., 2013), and consequently, inhibiting the activity of MAO-A by medications called monoamine oxidase inhibitors (MAOIs) is a well-established treatment approach in MDD. In a pilot study, Domschke et al. investigated the relationship between DNAm of the monoamine oxidase A (*MAO-A)* gene at 43 CpGs and escitalopram treatment response in a sample of 94, (61 females and 33 males)(Domschke et al., 2015). To account for *MAO-A* being encoded on the X chromosome, the authors stratified their analysis by sex. The authors did not observe any considerable influence of *MAO-A* methylation on treatment outcomes but hypomethylation at two CpGs in the *MAO-A* promoter region was nominally associated with a treatment response in females.

Tryptophan hydroxylase-2 methylation (*TPH2)*, another gene with important regulatory functions regarding serotonin synthesis has also been investigated recently (Shen et al., 2020). *TPH2* is involved in limiting the rate of serotonin synthesis by converting tryptophan to 5-hydroxytryptophan (Walther et al., 2003) and has been explored within the context of psychiatric disorders (Ottenhof, Sild, Levesque, Ruhe, & Booij, 2018), including MDD(Fan et al., 2021; S. J. Tsai et al., 2009; Zhang et al., 2015). Shen and colleagues (2020) assessed DNAm of *TPH2* in relation to antidepressant treatment response in 291 participants (female n=187, male n =104) with MDD. The authors stratified their analysis by sex and identified one hypermethylated CpG site in females and two in males to be nominally associated with an impaired treatment response. When assessing the impact of early life stress and DNAm on treatment outcomes, the authors observed childhood adversity to be correlated with hypomethylation of one CpG-site of *TPH2* in males, which remained associated with an impaired treatment response after FDR correction. Participants with high levels of childhood adversity had poorer treatment outcomes, leaving it unclear whether treatment resistance was mediated by differential methylation or childhood adversity.

In summary, studies assessing DNAm of monoamine-related loci in relation to treatment outcomes in MDD have been inconclusive and present significant methodological heterogeneity. The monoamine-related genes that are the focus of multiple independent studies, such as the serotonin transporter gene or its receptor subtypes, report inconsistent and even contradicting directions of effects associated with treatment outcomes. Interpretation of these efforts may be further complicated by assessing different epigenetic loci, as well as heterogeneity in treatment regimens and durations. These findings suggest future efforts are necessary in order to adequately assess the predictive potential of monoamine-related loci DNAm in MDD treatment outcomes.

#### 3.1.B Brain-derived neurotrophic factor (BDNF)

Another important pathway in MDD, and among the first ones to be examined in a pharmaco-epigenetic context with a treatment response (Tadic et al., 2014), is the neurotrophin Brain-derived neurotrophic factor (*BDNF). BDNF* has been extensively explored as a key molecular pathway in MDD involved in many synaptic and brain-related functions (Jin, Sun, Yang, Cui, & Xu, 2019), informing the neurotrophic hypothesis of MDD by tying the pathogenesis of depression to alterations in *BDNF* expression and functioning (Kozisek, Middlemas, & Bylund, 2008). Altered BDNF levels have been reported in patients with MDD in a number of studies (Bus et al., 2015; Kishi, Yoshimura, Ikuta, & Iwata, 2017). The expression of *BDNF* and Tropomyosin receptor kinase B *(TrkB)* has been identified to be critical for the action of several antidepressants (Casarotto et al., 2021; Castren & Monteggia, 2021) including SSRIs, TCAs, MAOIs, and ketamine, and alteration in the BDNF-TrkB multiprotein complex has been found to interfere with the effects of antidepressants (Casarotto et al., 2021). Additionally, *BDNF* plays a crucial role in other pathways implicated in MDD, due to its role downstream of the monoamine signaling pathway (Tayyab et al., 2018), additionally rendering *BDNF* an attractive target for pharmaco-epigenomic studies.

To date, six candidate gene studies have examined DNAm of *BDNF* in relation to antidepressant treatment response. Tadic and colleagues (2014) provided early preliminary evidence that depressed patients with hypomethylation in the promoter region of *BDNF* are less likely to respond to antidepressant treatment(Tadic et al., 2014). In a pilot study, the authors investigated DNAm at 13 CpGs of the *BDNF* promoter in the peripheral blood of 39 patients with MDD (female n= 22 male n = 17). DNAm was assessed using a targeted DNAm approach using next-generation sequencing of the bisulfite-converted DNA of the participants. Patients with lower methylation at CpG site 87 of exon IV had a significantly lower treatment response to fluoxetine and venlafaxine as compared to the participants showing higher methylation at that locus. Later attempts to validate such findings by the same group in a sample of 561 patients did not confirm this finding in the overall sample, however, a subgroup of 199 patients with severe depression (HAM-17 scores ≥ 25) documented significantly higher DNAm of that locus in treatment responders (Lieb et al., 2018). It’s noteworthy that the treatment regime of participants in the pilot study (Tadic et al., 2014) included a range of different antidepressants including SSRIs, SNRIs, and TCAs while the larger study (Lieb et al., 2018) only included patients who were administered escitalopram or venlafaxine.

These early findings of *BDNF* promoter methylation’s positive association with treatment outcomes have since been confirmed to hold potential in predicting antidepressant treatment response in similar investigations by other groups(Hsieh, Lin, Lee, & Huang, 2019; L. Li et al., 2021; Wang, Zhang, et al., 2018). A clinical trial by Wang and colleagues (Wang, Zhang, et al., 2018) investigated the association of *BDNF* gene methylation and genotype with antidepressant treatment response in 85 patients with MDD. After eight weeks’ treatment with the SSRI escitalopram, participants were assessed for changes in their depressive symptoms. Hypomethylation in promoter-region CpG islands at baseline was associated with an impaired treatment response compared to those who showed remission. Additionally, the authors document significantly better treatment outcomes in patients who had experienced fewer stressful life events along with having *BDNF* hypermethylation. Similarly, Li and colleagues (2021) examined the association of DNAm *BDNF* and response to antidepressant treatment in MDD in 291 patients. The authors interrogated a total of 194 CpG sites at baseline and observed significant hypermethylation of CpG 140 in the *BDNF* promotor in female MDD remitters but not in males (L. Li et al., 2021) warranting further examinations of sex-specific methylation patterns in relation to treatment response. Relatedly, a smaller study conducted in Taiwan observed hypermethylation at one CpG of the *BDNF* exon IX promoter was associated with response to antidepressant treatment in 25 responders(Hsieh et al., 2019). Conversely, a randomized control trial by Kim and colleagues documented the opposite direction of effect, in which *BDNF* exon VI hypermethylation was associated with treatment resistance to 24 week-long escitalopram administration, in 127 patients with comorbid acute coronary syndrome (J. M. Kim et al., 2015). However, the clinical heterogeneity among the patients with regard to cardiovascular pathologies and associated treatments limits the comparability of the findings to other pharmaco-epigenetics studies of MDD.

Collectively, despite notable heterogeneity in study designs, five out of six studies focused on *BDNF* converge on identifying a pattern of significant association between promoter methylation and response to antidepressant treatment. While hypomethylation of *BDNF* promoter CpGs was indicative of impaired treatment outcomes in two independent studies (Tadic et al., 2014; Wang, Zhang, et al., 2018) hypermethylation of this locus has been associated with positive antidepressant responses in three studies (Hsieh et al., 2019; L. Li et al., 2021; Lieb et al., 2018).

#### 3.1.C Interleukins

A large body of literature has characterized the role of inflammatory processes in MDD (reviewed in (Beurel, Toups, & Nemeroff, 2020; Pariante, 2017)). Immune cell-mediated inflammation is a key mechanism to maintain homeostasis in the response to harmful stimuli such as pathogens or cell damage and prolonged inflammatory immune responses have been linked to MDD (Beurel et al., 2020). A range of proinflammatory cytokines and their corresponding receptors have been associated with MDD (Petralia et al., 2020) including Interleukin (IL)-6, IL, IL-1β, IL-2, IL-4, IL-10, the IL-1, tumor necrosis factor (TNF)-α, transforming growth factor-β, and also C-reactive protein. Many mechanisms contributing to inflammation in MDD have been proposed. These include the inflammasome pathway activated by heightened levels of damage-associated molecular patterns (Fleshner, Frank, & Maier, 2017), oxidative stress (Bajpai, Verma, Srivastava, & Srivastava, 2014), psychosocial stress (I. B. Kim, Lee, & Park, 2022), and diet (Lassale et al., 2019; Luo et al., 2023). Additionally, evidence from clinical trials further highlights the role of immune dysregulation in MDD (Bai et al., 2020; Wittenberg et al., 2020). Finally, the immune system’s interrelation with the neuroendocrine system makes this pathway particularly relevant to MDD etiology and an important target for epigenetic treatment response prediction in MDD.

For example, an early randomized controlled trial by Powell and colleagues (2013) (Powell et al., 2013) investigated DNAm of the *interleukin-11* (*IL11*) as a general predictor of antidepressant treatment response in the peripheral blood of a sub-sample of 113 (female n= 67 male n = 46) participants with MDD. The patients were treated with escitalopram (n=80) or the tricyclic antidepressant nortriptyline (n= 33) for 12 weeks. Their analysis interrogated one CpG island, a region with a high number of CpG dinucleotide repeats, in *IL11* using a mass spectrometry approach. Their analysis revealed that hypomethylation of *IL11* CpG unit 5 was associated with an increased response to both administered treatments. Linear regression analysis assessing the predictive potential of DNAm at any of the assessed CpGs revealed an interesting contrast in the direction of the association of DNAm and treatment response: while increased levels of baseline DNAm at *IL11* unit 4 was found to be associated with a higher treatment response among participants taking escitalopram, an impaired response was observed in those taking nortriptyline, pointing to medication-specific difference. Such medication specific associations make a strong case to, if study design permits, stratify future analysis in pharmaco-epigenetic studies by different antidepressant treatments. Additionally, the authors report that patients with higher *IL11* methylation in combination with the *rs1126757* allele had better antidepressant responses by the end of treatment, providing evidence of how assessment of DNAm and genotype can be combined to predict treatment outcomes in MDD.

Another recent pharmacogenetics study by Draganov *et al*., 2019 (Draganov et al., 2019), investigated if the response to antidepressant treatment is associated with DNAm of several inflammation-related genes. Similarly, they assessed if a treatment response is associated with having inflammation-specific genetic variation (SNPs). The authors identified 41 SNPs located in inflammation regulatory genes including *interleukin 1-β, interleukin 2, 6, IL6R, IL10, IL18*, tumor necrosis factor *(TNF)-α*, and interferon *(IFN)-γ*. Subsequent pyrosequencing of five selected CpG Islands within *IL1-β*, *IL6,* and *IL6R* quantified the methylation status and revealed a higher methylation percentage of *IL6R* CpG island to be associated with treatment response. SNPs in *IL1-β* and, marginally, *IL6R* were found to be in association with the treatment response to a range of treatments including SSRIs, TCAs, MAOIs, mood stabilizers, and antipsychotics. However, 56 of the patients were diagnosed with a range of comorbid mental disorders including affective, anxiety, psychotic, and schizo-affective disorder at baseline leaving the study findings to be interpreted with caution. In addition, the authors did not control for heterogeneity in antidepressant treatment types and dosages, further limiting the interpretability of the findings. In sum, findings from the two candidate studies investigating interleukins are largely inconclusive, only nominally significant (Draganov et al., 2019), and show significant heterogeneity in study design.

#### 3.1.D Summary of Challenges associated with Candidate Gene Studies to Date

Apart from the limited number of studies overall, there are several reasons complicating the consistency between findings of candidate gene studies to date, including: (1) The scope of assessed CpGs differs greatly. For example, in the above-described studies of *SLC6A4,* the number of CpGs investigated range between 7(Kang et al., 2013) and 81 (Okada et al., 2014), with only Kang et al (Kang et al., 2013) and Domschke *et al (*Domschke et al., 2014) and Schiele *et al*., 2021 (Schiele et al., 2021) examining some (n=7) of the same CpGs within that locus. Similarly, there has been scarce convergence regarding assessed CpGs in studies assessing *BDNF* or serotonin receptor subtypes, with only two studies (respectively, (Lieb et al., 2018; Tadic et al., 2014) & (Gao et al., 2022; Xu et al., 2022)) interrogating the same CpGs (2) The pharmacological intervention in the studies is quite heterogeneous with only few studies focusing on one SSRI alone(Wang, Lv, et al., 2018; Wang, Zhang, et al., 2018). Some studies augment SSRI treatment with antipsychotics, benzodiazepines and mood stabilizers(Domschke et al., 2014; Gasso et al., 2017) while others incorporate a range of SSRIs, SNRIs, and TCAs(L. Li et al., 2021; Okada et al., 2014; Schiele et al., 2019). Despite acknowledging the heterogeneity in medications among the patients, only a limited number of studies stratify their analysis by different antidepressants(Domschke et al., 2015; Powell et al., 2013; Xu et al., 2022) (3) Differences in allele frequencies of assessed candidate loci. For example, *SLC6A4* allele frequency differences(Ng et al., 2006) between Asian (Iga et al., 2016; Kang et al., 2013; Moon, Kim, Kim, Lim, & Kim, 2023; Okada et al., 2014) and Europeans (Domschke et al., 2014; Schiele et al., 2021) study populations can also affect the observed methylation status (Giri et al., 2017) (4) The treatment duration varies significantly. While the shortest treatment duration was about 2 weeks (Gao et al., 2022) the longest treatment course was up to 24 weeks(J. M. Kim et al., 2015). The vast variation in treatment duration is also observed among studies using a similar inventory of SSRIs, SNRIs, and TCAs, NaSSAs (Kang et al., 2013; L. Li et al., 2021). (5) The majority of studies fail to address the potential confounding effect of blood cell type heterogeneity. Only one recent candidate study (Shen et al., 2020) included white blood cell proportion as a covariate in their analysis while the other studies allowed the potential of observing inaccurate methylation patterns due to differences in cell-type compositions in the blood samples of the participants. Of note, many of these issues (2-5) apply not just to candidate gene studies, but also to genome-scale studies, described in more detail below. Taken together, these shortcomings illustrate the difficulty of consolidating findings from existing candidate genes studies, rendering an assessment of their utility as biomarkers with predictive potential likely premature.

### 3.3 Epigenome-wide association studies of response to antidepressants

Genome-scale studies have the potential to reveal novel loci and pathways associated with MDD that have not been considered before in candidate gene approaches. To date, only six studies have used a genome-scale approach to elucidate the relationship between baseline DNAm and treatment outcomes in MDD of which four efforts have been pharmaco-epigenetic studies (Engelmann et al., 2022; Ju et al., 2019; Martinez-Pinteno et al., 2021; Takeuchi et al., 2017).

In a study from Japan, Takeuchi and Colleagues (2018) (Takeuchi et al., 2017) investigated the potential of predicting the treatment response to paroxetine based on genome-scale DNAm patterns in the peripheral blood of 68 MDD patients. In their analysis, the authors contrasted the ten best responders and the ten worst responders to paroxetine treatment and identified the hypermethylation of the genes liprin-alpha-4 (*PPFIA4)* and heparan sulfate-glucosamine 3-sulfotransferase 1 (*HS3ST1)* at baseline in patients with the worst treatment outcomes in relation to the best treatment responders (Takeuchi et al., 2017). The limited sample size of their study, not adjusting for cell-type heterogeneity, along with the analytic approach of focusing exclusively on selected best and worst responders to paroxetine treatment pose challenges to the interpretability of the findings as not every patient falls among the extremes of the spectrum. Ju and colleagues investigated global DNAm changes in the blood of responders (*N* = 82) and non-responder (*N* = 95) to the eight-week-long escitalopram treatment of 177 patients with MDD(Ju et al., 2019). Their analysis revealed 303 differentially methylated positions (DMPs) between treatment responders and non-responders at baseline at a nominal p-value (p < 0.05) with several differentially methylated CpG-site-associated differences in gene expression between the assessed groups. One hypomethylated position in *CHN2* showed the most significant association with mRNA expression and was replicated in an external cohort.

Another small preliminary study of adolescents with MDD (Martinez-Pinteno et al., 2021) assessed baseline DNAm differences in responders (*N* = 11) and non-responders (*N* = 11) before undergoing eight weeks of fluoxetine treatment. Considering a difference in β-value medians of 0.2 the authors identified 21 DMPs between treatment responders and the treatment-resistant group at baseline, predicting treatment outcomes for MDD. Their small sample size limits the interpretability of the findings; however, their data could be pooled together with future efforts.

A recent study profiled genome-scale baseline DNAm in responders and non-responders to escitalopram, venlafaxine, and lithium treatment in 80 European MDD patients (Engelmann et al., 2022). Despite the inability to identify any differentially methylated positions on an epigenome-scale associated with treatment response, the authors observed 18 hypermethylated and 2 hypomethylated regions associated with positive treatment outcomes including one site in *SORBS2* and *CYP2C18*.

Taken together, findings from genome-scale studies suggest that using hypothesis-free approaches may be promising for identifying DNAm based interindividual differences discerning treatment responders and non-responders.

### 3.4 Brain stimulation therapies (ECT, TMS)

Another alternative non-pharmacological treatment for MDD, which is now more investigated for potential DNAm changes associated with treatment response, is brain stimulation therapies such as ECT and TMS. TMS treatment involves stimulating the brain via a magnetic field through the intact scalp. This exposure results in acute depolarization and altered electrical activity in the underlying neurons; when applied repetitively, TMS alters the excitability of the surrounding region for a sustained period outlasting the TMS exposure and is thought to exert its therapeutic effects through altered cortical excitability (Jannati, Oberman, Rotenberg, & Pascual-Leone, 2023). To date, we have not identified any published studies examining the epigenetic profiles of responders to TMS treatment however, a few studies (*N* = 5) have examined blood-derived DNAm profiles in relation to response to ECT in MDD. ECT treatment is a procedure in which electric currents are applied to the brain resulting in brief and controlled generalized seizures (M. Li et al., 2020). Although the mechanism of action underlying ECT treatment remains elusive, findings from previous research converge on attributing its therapeutic effects to augmentations in neuroplasticity (Deng et al., 2022; Ousdal et al., 2022).

The first ECT-related study by Kleimann and colleagues (Kleimann et al., 2015) reported lower levels of *BDNF* promoter I, IV, and VI CpGs in the peripheral blood of 11 MDD patients who remitted following treatment using targeted bisulfite-pyrosequencing. The authors identified a negative correlation between mean *BDNF* promoter methylation levels in the blood and BDNF serum levels which have previously been observed by others (Boulle et al., 2012). A more recent study by Moshny and Colleagues(Moschny, Jahn, et al., 2020) investigated DNAm of two genes involved in the production of brain-derived neurotrophic factor (BDNF), *t-PA* and *PAI-1*, as potential biomarkers of ECT responsiveness when measured in peripheral blood; however, no differences in baseline *t-PA* or *PAI-1* DNAm levels were detected between eventual ECT responders vs. non-responders in either their first cohort (n=58) or in their second cohort (n=28). Another preliminary study by the same research group (Moschny, Zindler, et al., 2020) assessed genome-scale DNAm in peripheral blood mononuclear cells longitudinally in a small (n=12) cohort of patients with MDD, identifying eight novel candidate genes implicated in the response to ECT treatment after analyzing the variance of every differentially methylated probe including *RNF175, RNF213, TBC1D14, TMC5, WSCD1 AC018685.2, AC098617.1, CLCN3P1*. Further, the DNAm of two CpG sites within the genes *AQP10* and *TRERF1* were found to change during treatment.

An ECT study by Sirignano and colleagues (Sirignano et al., 2021) examined the effects of ECT treatment on DNAm associated with response status in a small sample of depressed patients (n = 34). The authors assessed DNAm at baseline and also post completion of the treatment. In the binary response model of responders vs. non-responder status, one genome-scale differentially methylated CpG site was identified in *TNKS*, a protein-coding gene involved in biological processes such as the Wnt signaling pathway and telomere length, both of which have previously been implicated in MDD (Ridout, Ridout, Price, Sen, & Tyrka, 2016; Tayyab et al., 2018). Additionally, the author identified two differentially methylated regions associated with reduction in depression measures. Subsequently, the authors investigated CpG sites in candidate genes (n=799), previously implicated in the literature to be involved with DNAm in antidepressant medication. However, their analysis of candidate CpGs associated with treatment ECT response revealed only one hypermethylated CpG in *FKBP5* to be significant after multiple test corrections in their continuous response model.

Taken together, the few studies examining genome-scale DNAm patterns in association with response to ECT treatment are too scarce to infer any trends among the suggested differentially methylated loci leaving plenty of opportunities for future studies. Many of the identified loci, from the above-mentioned preliminary efforts, require replication in more extensive cohorts of patients with MDD.

## 4. Discussion

The objective of this review was to synthesize existing literature characterizing DNAm patterns associated with treatment outcomes of patients undergoing various forms of treatment for MDD. The literature reviewed included several candidate gene and genome-scale studies examining different treatment modalities for MDD including various antidepressants and ECT. Overall, epigenetic studies investigating DNAm associated with treatment responses are in their infancy with a limited number of studies that are frequently based on small sample sizes with inconsistent finding across studies. Significant heterogeneity exists in study design, evaluated loci, tissue type examined, methylation profiling approaches, treatment duration, dosage and type paired with other patient clinical characteristics, rendering premature any conclusions about the clinical utility of implicated loci. The scarcity of existing studies and the inconsistent nature of the results across studies suggests the need for more research into DNAm patterns associated with treatment outcomes; this is particularly true for non-pharmacological approaches including brain stimulation therapies (e.g., TMS and ECT) and psychotherapy, for which there remains a dearth of information in relation to response to those treatments in MDD. Limited evidence from a study of children with anxiety disorders undergoing cognitive behavior theory (Roberts et al., 2014) observed changes in *SLC6A4 to* be associated with treatment response, which gives hope for efforts to characterize other treatment modalities that will follow.

Despite the lack of major convergence of differentially methylated loci among most studies evaluating DNAm in relation to treatment response to MDD treatments, *BDNF* perhaps emerges to be a notable exception. Hypomethylation of the *BDNF* promoter has been implicated to be predictive of treatment non-response in two studies with MDD patients (Tadic et al., 2014; Wang, Zhang, et al., 2018) while hypermethylation of this locus has been associated with treatment success in three studies(Hsieh et al., 2019; L. Li et al., 2021) including one study with patients suffering from severe MDD (Lieb et al., 2018). Interestingly, the positive association between *BDNF* promoter methylation and treatment responses has only been observed in pharmacological treatment approaches. The only non-pharmacological study with patients undergoing ECT treatment (Kleimann et al., 2015) observed the hypomethylation of *BDNF* promoter CpGs among MDD remitters. Lastly, the finding of Li and colleagues reporting significant hypermethylation of CpGs in *BDNF* in female MDD remitters, but not in males, warrants further investigations into sex-specific DNAm trends in relation to treatment response.

Another important finding from those initial studies is reports of medication-specific differences in regards to DNAm associated with treatment response. Powel and colleagues (2013) identified increased levels of baseline DNAm at *IL11* unit 4 associated with a higher treatment response among participants taking escitalopram but an impaired response in those on nortriptyline therapy (Powell et al., 2013). If replicated by additional independent studies, such findings show the potential of pharmaco-epigenetic studies in informing future treatment decisions, while also highlighting the need for more comprehensive future studies that stratify patients into groups according to their treatment, if various antidepressants are utilized.

This literature review revealed several challenges and methodological considerations, that might hinder progress on the successful identification of predictive, DNAm-based biomarkers with clinical utility. Chief among such challenges is the relative instability of epigenetic marks in comparison to genetic variation. For example, genetic assays like GWAS measure allele frequencies of genetic variants among study participants that are consistent across various cell types and stable over time. On the other hand, epigenetic marks such as DNA methylation are dynamic and susceptible to changes mediated by a range of environmental factors including aging, psychological stress, and importantly, antidepressant medication (Moon et al., 2023; Wang, Zhang, et al., 2018) and ECT (Moschny, Jahn, et al., 2020; Schurgers et al., 2022; Sirignano et al., 2021). Several studies document mechanisms in which psychotropic medications can alter the epigenome, making controlling for such alteration an important consideration (Bayles et al., 2013; Gassen et al., 2015). For example, with regard to DNAm, Gassen *et al*. demonstrated that the administration of paroxetine reduces the phosphorylation and activity of DNA methyltransferase 1 in the peripheral blood of patients with MDD(Gassen et al., 2015). Evidence from cell culture and murine studies suggests additional mechanisms in which antidepressant medications can influence the epigenome by alterations to methyl-CpG-binding protein domains (Csoka & Szyf, 2009) or chromatin remodelers (Qiao et al., 2019) while emerging literature documents the effects of antidepressant treatments on the methylome (Mohammadi et al., 2022; Moon et al., 2023; Wang, Zhang, et al., 2018). Similarly, emerging longitudinal ECT DNAm studies observed alterations in DNAm in patients undergoing ECT (Moschny, Zindler, et al., 2020; Schurgers et al., 2022; Sirignano et al., 2021). Overcoming these challenges requires carefully designed longitudinal efforts that ideally assess epigenetic marks at multiple timepoints.

Another substantial challenge in the development of biomarkers that could predict treatment responses based on DNAm is the significant heterogeneity in cell-type specific DNAm (Loyfer et al., 2023), making the selection of the tissue in which such biomarkers are assessed an important consideration. This is particularly true for the assessment of psychiatric disorders like MDD, for which the organ central to the disease, the brain, is not accessible in living patients. Therefore, investigators are limited to peripheral tissues such as blood, saliva, or buccal tissue. Despite some progress in the characterization of the concordance between epigenetic alteration observed in blood and in the brain (Braun et al., 2019; Hannon, Lunnon, Schalkwyk, & Mill, 2015; Loyfer et al., 2023), the question of which tissue is most reflective of the brain remains to be answered by future studies. To overcome this challenge, investigators could focus their efforts on CpGs with a high concordance between the peripheral tissue of choice and the brain, informed by consulting publicly available resources such as the Epigenomics Roadmap Consortium (Roadmap Epigenomics et al., 2015), the PsychENCODE knowledge portal (Psych et al., 2015) or web tools such as BECon (Edgar, Jones, Meaney, Turecki, & Kobor, 2017), Image-CpG (Braun et al., 2019) or wgbstools (Loyfer et al., 2023). All studies identified in this review, and the vast majority of epigenetic studies investigating DNAm in general, have been conducted in peripheral blood.

Another important consideration is the limited scope of most DNAm treatment response studies to date. The majority of conducted studies are candidate-gene studies examining loci such as *SLC6A4*, *BDNF*, *IL-1*, *IL-6* & *IL-11, MAOA, HTR1A/1B,* and *THH2.* Although the investigated genes to date are central to many important pathways in depression such as monoamines, neurotrophins, and inflammation, it appears that such pharmaco-epigenetic studies of the stress pathway are less represented. Figure 4 illustrates the genes and pathways assessed in extant candidate studies investigating DNAm in relation to treatment responses. We identified only one candidate gene study (Maier et al., 2023) focused on loci of the stress pathway in the context of DNAm studies investigating response to MDD treatments, despite stress being a well-studied pathway in MDD pathophysiology. Stress exposure, especially during developmentally critical periods (Heim & Binder, 2012; Klengel et al., 2013), is the most well-characterized risk factor in MDD (Z. Li et al., 2021) and has also been extensively studied in the context of epigenetics (Penner-Goeke & Binder, 2019). Stress exposure has been reported to induce structural changes in areas of the brain central to MDD (McEwen, Nasca, & Gray, 2016) making the investigation of such loci in relation to their association with a treatment response a plausible target for candidate gene studies. One of those loci could be FK506 binding protein 5 (*FKBP5*), an important regulator of the HPA-axis crucial to the termination of the stress response by a negative feedback loop. Genomic variations of this locus have previously been associated with HPA-axis alterations, recurrence of symptoms as well as response to treatment (Binder et al., 2004). Additionally, findings from cell-culture and animal studies, demonstrate the breath of mechanism in which *FKBP5* exerts its effect on this locus with other stress-related pathways including BDNF (Gassen et al., 2015) and the immune system(Annett, Moore, & Robson, 2020). Other genes that could serve as a potential target for pharmaco-epigenetic studies focused on treatment response could be Corticotropin-releasing factor-binding protein *(CRHB)* or the glucocorticoid receptor (*NR3C1)* due to their roles in the stress pathway and evidence from previous clinical studies (Humphreys et al., 2019; Roy, Dunbar, Shelton, & Dwivedi, 2017).

**Figure 4.**
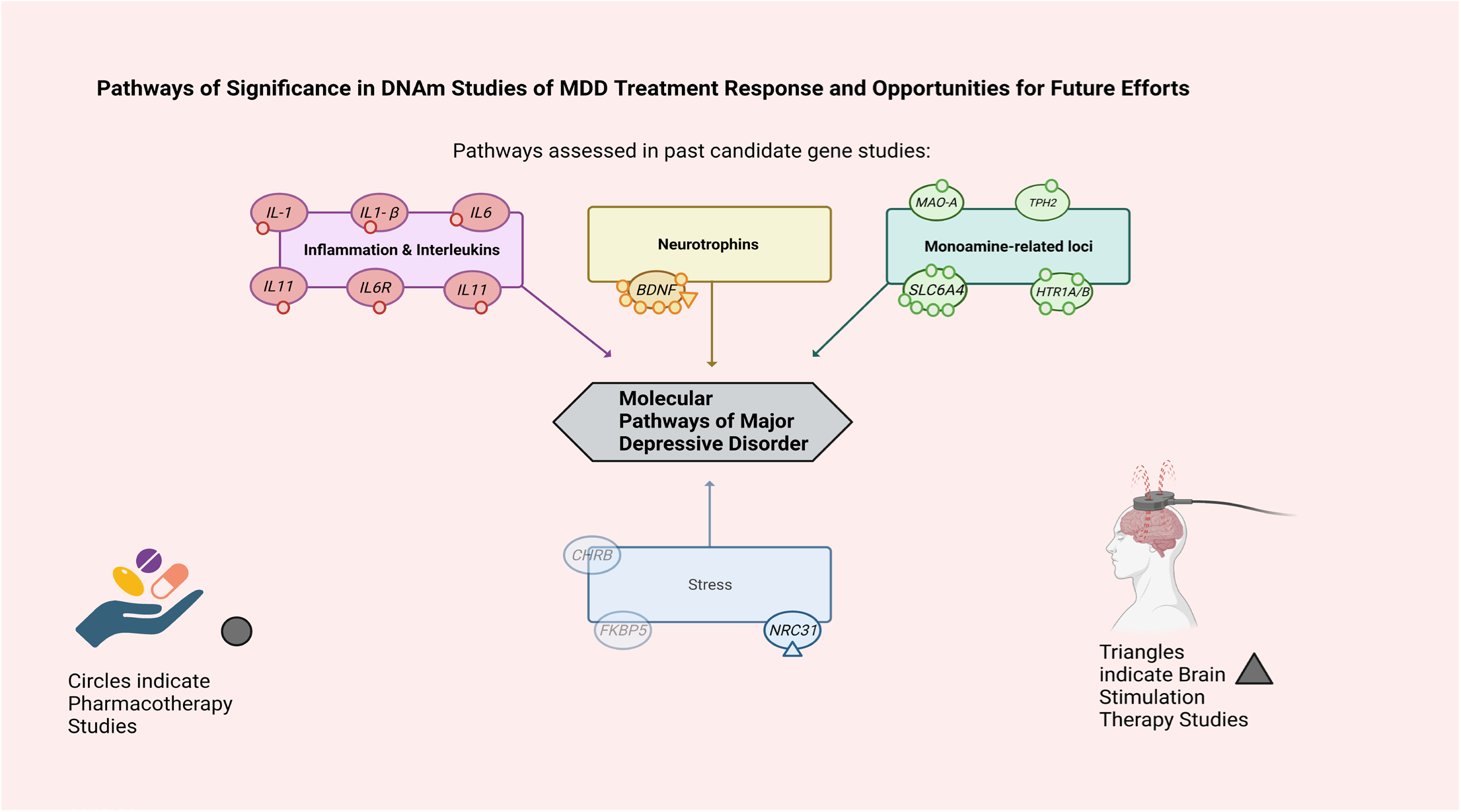
Pathways of Significance in Pharmaco-epigenetic Studies of MDD Treatment Response and Opportunities for Future Efforts.

Despite such opportunities for candidate gene studies, the application of genome-scale approaches will be more appropriate to elucidate potentially new biological pathways associated with MDD. Epigenome-wide association studies (EWAS) could reveal novel epigenetic loci involved in treatment response. Smaller studies could benefit from larger consortia efforts that combine data to have enough power to detect epigenome-scale differences (P. C. Tsai & Bell, 2015). Another important consideration for genome-scale approaches is the heterogeneity in technologies used. While one study (Takeuchi et al., 2017) used older array technology assessing 450,000 CpGs, four studies employed the Epic Methylation array capable of interrogating 850,000 probes(Engelmann et al., 2022; Ju et al., 2019; Martinez-Pinteno et al., 2021; Sirignano et al., 2021). Moshny and Colleagues used the Nextseq 550 with EPIC kit capable of interrogating over 3.3 million CpGs (Moschny, Zindler, et al., 2020). Such differences in the scope of the interrogated CpGs can make it difficult to compare findings, especially among loci that are not covered by both of the above-mentioned microarrays. Furthermore, the number of CpGs covered by current Illumina BeadChip platforms only accounts for around 3% of all GpG sites in the human genome (Bibikova et al., 2011) leaving ample opportunities for future studies using whole-genome bisulfite sequencing (WGBS).

An additional limitation that could potentially be addressed in the future by WGBS studies is that studies to date have exclusively focused on DNAm at CpG dinucleotides. Compelling evidence reports on the wide distribution of cytosine methylation preceding adenine, thymine, or other cytosine (non-CpG methylation) in several brain tissues (de Mendoza et al., 2021) and neurons with non-CpG methylation accounting for as much as 25% of the methylation observed in neurons (Guo et al., 2014). WGBS studies offer opportunities to identify methylation patterns important in the pathogenesis and treatment response of MDD in previously unexplored loci including non-CpG methylation.

## 5. Conclusion

It is evident that pharmaco-epigenetic studies characterizing DNAm patterns associated with treatment outcomes in MDD are in an early stage. Many efforts suffer from insufficient statistical power due to their limited sample sizes hindering the detection of biomarkers with predictive utility. The majority of findings from previous studies have yet to be replicated in independent cohorts suggesting that well-designed studies investigating the DNAm associated with a treatment response have much to contribute. Those studies will be most informative when multiple genomic measures are integrated into epigenetic studies of depression (Engelmann et al., 2022; Fuh, Fiori, Turecki, Nagy, & Li, 2023; Ju et al., 2019; Wang, Lv, et al., 2018; Wang, Zhang, et al., 2018), in order to gain more wholistic insights into the functional relevance of observed epigenetic variation in MDD treatment response. Assessing DNAm at multiple timepoints throughout the treatment could be particularly informative and perhaps bring clinically significant opportunities. For example, DNAm profiled early in the course of a new intervention could elucidate methylation signatures that are associated with an early treatment response or treatment resistance, and in turn, inform the likelihood of remission at the end of the treatment. This would enable clinicians to be make timely adjustments, sparing valuable time for patients, reduce suffering, mitigate the burden of side effects, and cut healthcare expenditures.

In the future, well-designed studies may help elucidate additional biomarkers that can predict MDD treatment response and potentially inform treatment options for a patient suffering from treatment-resistant depression. The considerable cost of conducting controlled clinical trials, environmental effects on the epigenome and inter-individual epigenomic variation pose challenges when it comes to efforts to reproduce findings in larger cohorts. Findings from smaller studies should be considered preliminary and should be replicated in larger cohorts. Larger collaborative efforts including several sites are key to overcoming sample size limitations. Previous efforts such as the STAR*D project (Rush et al., 2004) are a good example of such collaborative efforts. Additionally, there is increasing evidence that using predictive models based on multimodal data fares much better in the prediction of treatment outcomes than low-dimensional data (Gao et al., 2022; Lee et al., 2018). In this regard, future efforts should integrate genetic information with epigenetic approaches to identify more reliable prediction models. Approaches akin to the ones applied by Wang and colleagues (Wang, Zhang, et al., 2018) in which life stress measures combined with DNAm gave the best prediction of treatment response, or Powell and colleagues reporting the interaction of *IL11* methylation and the rs1126757 variation (Powell et al., 2013) are great examples of how multiple genetic, epigenetic and psychosocial predictors can be combined to inform future response prediction tools. Similarly, multi-omics study designs like the one employed by Ju and colleagues (Ju et al., 2019) or Schurgers *et al*. (Schurgers et al., 2022), integrating DNAm measures with transcriptomic data(Ju et al., 2019; Schurgers et al., 2022), can provide valuable functional insights of MDD pathophysiology and inform about interplay between gene expression and DNAm. Lastly, future efforts could expand on treatments other than antidepressants and ECT by investigating DNAm in treatment responders undergoing other forms of treatments such as TMS or psychotherapy, while also including participants of ancestries beyond Europeans and Asians.

## Data Availability

All data produced in the present work are contained in the manuscript

## Acknowledgments and Disclosures

J. Dahrendorff was supported by the College of Public Health Doctoral Fellowship of the University of South Florida. We would like to thank Allison Howard for her instructions on effective literature searching for the students in the College of Public Health. Figures 1 and 4 created with BioRender (www.biorender.com)

## References

1. Andersson, N. W., Gustafsson, L. N., Okkels, N., Taha, F., Cole, S. W., Munk-Jorgensen, P., & Goodwin, R. D. (2015). Depression and the risk of autoimmune disease: a nationally representative, prospective longitudinal study. Psychol Med, 45(16), 3559–3569. doi:10.1017/S0033291715001488

2. Annett, S., Moore, G., & Robson, T. (2020). FK506 binding proteins and inflammation related signalling pathways; basic biology, current status and future prospects for pharmacological intervention. Pharmacol Ther, 215, 107623. doi:10.1016/j.pharmthera.2020.107623

3. Arnaud, A. M., Brister, T. S., Duckworth, K., Foxworth, P., Fulwider, T., Suthoff, E. D., … Reinhart, M. L. (2022). Impact of Major Depressive Disorder on Comorbidities: A Systematic Literature Review. J Clin Psychiatry, 83(6). doi:10.4088/JCP.21r14328

4. Association, A. P. (2013). Diagnostic and Statistical Manual of Mental Disorders (5th ed.). Washington, DC.

5. Bai, S., Guo, W., Feng, Y., Deng, H., Li, G., Nie, H., … Tang, Z. (2020). Efficacy and safety of anti-inflammatory agents for the treatment of major depressive disorder: a systematic review and meta-analysis of randomised controlled trials. J Neurol Neurosurg Psychiatry, 91(1), 21–32. doi:10.1136/jnnp-2019-320912

6. Bajpai, A., Verma, A. K., Srivastava, M., & Srivastava, R. (2014). Oxidative stress and major depression. J Clin Diagn Res, 8(12), CC04-07. doi:10.7860/JCDR/2014/10258.5292

7. Bayles, R., Baker, E. K., Jowett, J. B., Barton, D., Esler, M., El-Osta, A., & Lambert, G. (2013). Methylation of the SLC6a2 gene promoter in major depression and panic disorder. PLoS One, 8(12), e83223. doi:10.1371/journal.pone.0083223

8. Beurel, E., Toups, M., & Nemeroff, C. B. (2020). The Bidirectional Relationship of Depression and Inflammation: Double Trouble. Neuron, 107(2), 234–256. doi:10.1016/j.neuron.2020.06.002

9. Bibikova, M., Barnes, B., Tsan, C., Ho, V., Klotzle, B., Le, J. M., … Shen, R. (2011). High density DNA methylation array with single CpG site resolution. Genomics, 98(4), 288–295. doi:10.1016/j.ygeno.2011.07.007

10. Binder, E. B., Salyakina, D., Lichtner, P., Wochnik, G. M., Ising, M., Putz, B., … Muller-Myhsok, B. (2004). Polymorphisms in FKBP5 are associated with increased recurrence of depressive episodes and rapid response to antidepressant treatment. Nat Genet, 36(12), 1319–1325. doi:10.1038/ng1479

11. Bjornsson, H. T., Sigurdsson, M. I., Fallin, M. D., Irizarry, R. A., Aspelund, T., Cui, H., … Feinberg, A. P. (2008). Intra-individual change over time in DNA methylation with familial clustering. JAMA, 299(24), 2877–2883. doi:10.1001/jama.299.24.2877

12. Braun, P. R., Han, S., Hing, B., Nagahama, Y., Gaul, L. N., Heinzman, J. T., … Shinozaki, G. (2019). Genome-wide DNA methylation comparison between live human brain and peripheral tissues within individuals. Transl Psychiatry, 9(1), 47. doi:10.1038/s41398-019-0376-y

13. Breitling, L. P., Yang, R., Korn, B., Burwinkel, B., & Brenner, H. (2011). Tobacco-smoking-related differential DNA methylation: 27K discovery and replication. Am J Hum Genet, 88(4), 450–457. doi:10.1016/j.ajhg.2011.03.003

14. Brunner, E. J., Shipley, M. J., Britton, A. R., Stansfeld, S. A., Heuschmann, P. U., Rudd, A. G., … Kivimaki, M. (2014). Depressive disorder, coronary heart disease, and stroke: dose-response and reverse causation effects in the Whitehall II cohort study. Eur J Prev Cardiol, 21(3), 340–346. doi:10.1177/2047487314520785

15. Bus, B. A., Molendijk, M. L., Tendolkar, I., Penninx, B. W., Prickaerts, J., Elzinga, B. M., & Voshaar, R. C. (2015). Chronic depression is associated with a pronounced decrease in serum brain-derived neurotrophic factor over time. Mol Psychiatry, 20(5), 602–608. doi:10.1038/mp.2014.83

16. Casarotto, P. C., Girych, M., Fred, S. M., Kovaleva, V., Moliner, R., Enkavi, G., … Castren, E. (2021). Antidepressant drugs act by directly binding to TRKB neurotrophin receptors. Cell, 184(5), 1299–1313 e1219. doi:10.1016/j.cell.2021.01.034

17. Castren, E., & Monteggia, L. M. (2021). Brain-Derived Neurotrophic Factor Signaling in Depression and Antidepressant Action. Biol Psychiatry, 90(2), 128–136. doi:10.1016/j.biopsych.2021.05.008

18. Cecil, C. A. M., Zhang, Y., & Nolte, T. (2020). Childhood maltreatment and DNA methylation: A systematic review. Neurosci Biobehav Rev, 112, 392–409. doi:10.1016/j.neubiorev.2020.02.019

19. Chang, Z., Lichtenstein, P., Asherson, P. J., & Larsson, H. (2013). Developmental twin study of attention problems: high heritabilities throughout development. JAMA Psychiatry, 70(3), 311–318. doi:10.1001/jamapsychiatry.2013.287

20. Chilmonczyk, Z., Bojarski, A. J., Pilc, A., & Sylte, I. (2015). Functional Selectivity and Antidepressant Activity of Serotonin 1A Receptor Ligands. Int J Mol Sci, 16(8), 18474–18506. doi:10.3390/ijms160818474

21. Coppen, A. (1967). The biochemistry of affective disorders. Br J Psychiatry, 113(504), 1237–1264. doi:10.1192/bjp.113.504.1237

22. Csoka, A. B., & Szyf, M. (2009). Epigenetic side-effects of common pharmaceuticals: a potential new field in medicine and pharmacology. Med Hypotheses, 73(5), 770–780. doi:10.1016/j.mehy.2008.10.039

23. de Mendoza, A., Poppe, D., Buckberry, S., Pflueger, J., Albertin, C. B., Daish, T., … Lister, R. (2021). The emergence of the brain non-CpG methylation system in vertebrates. Nat Ecol Evol, 5(3), 369–378. doi:10.1038/s41559-020-01371-2

24. Deng, Z. D., Argyelan, M., Miller, J., Quinn, D. K., Lloyd, M., Jones, T. R., … Abbott, C. C. (2022). Electroconvulsive therapy, electric field, neuroplasticity, and clinical outcomes. Mol Psychiatry, 27(3), 1676–1682. doi:10.1038/s41380-021-01380-y

25. Diniz, B. S., Butters, M. A., Albert, S. M., Dew, M. A., & Reynolds, C. F., 3rd. (2013). Late-life depression and risk of vascular dementia and Alzheimer’s disease: systematic review and meta-analysis of community-based cohort studies. Br J Psychiatry, 202(5), 329–335. doi:10.1192/bjp.bp.112.118307

26. Domschke, K., Tidow, N., Schwarte, K., Deckert, J., Lesch, K. P., Arolt, V., … Baune, B. T. (2014). Serotonin transporter gene hypomethylation predicts impaired antidepressant treatment response. Int J Neuropsychopharmacol, 17(8), 1167–1176. doi:10.1017/S146114571400039X

27. Domschke, K., Tidow, N., Schwarte, K., Ziegler, C., Lesch, K. P., Deckert, J., … Baune, B. T. (2015). Pharmacoepigenetics of depression: no major influence of MAO-A DNA methylation on treatment response. J Neural Transm (Vienna), 122(1), 99–108. doi:10.1007/s00702-014-1227-x

28. Draganov, M., Arranz, M. J., Salazar, J., de Diego-Adelino, J., Gallego-Fabrega, C., Jubero, M., … Portella, M. J. (2019). Association study of polymorphisms within inflammatory genes and methylation status in treatment response in major depression. Eur Psychiatry, 60, 7–13. doi:10.1016/j.eurpsy.2019.05.003

29. Edgar, R. D., Jones, M. J., Meaney, M. J., Turecki, G., & Kobor, M. S. (2017). BECon: a tool for interpreting DNA methylation findings from blood in the context of brain. Transl Psychiatry, 7(8), e1187. doi:10.1038/tp.2017.171

30. Engelmann, J., Zillich, L., Frank, J., Wagner, S., Cetin, M., Herzog, D. P., … Streit, F. (2022). Epigenetic signatures in antidepressant treatment response: a methylome-wide association study in the EMC trial. Transl Psychiatry, 12(1), 268. doi:10.1038/s41398-022-02032-7

31. Fakhoury, M. (2015). New insights into the neurobiological mechanisms of major depressive disorders. Gen Hosp Psychiatry, 37(2), 172–177. doi:10.1016/j.genhosppsych.2015.01.005

32. Fan, R., Hua, T., Shen, T., Jiao, Z., Yue, Q., Chen, B., & Xu, Z. (2021). Identifying patients with major depressive disorder based on tryptophan hydroxylase-2 methylation using machine learning algorithms. Psychiatry Res, 306, 114258. doi:10.1016/j.psychres.2021.114258

33. Fleshner, M., Frank, M., & Maier, S. F. (2017). Danger Signals and Inflammasomes: Stress-Evoked Sterile Inflammation in Mood Disorders. Neuropsychopharmacology, 42(1), 36–45. doi:10.1038/npp.2016.125

34. Flint, J. (2023). The genetic basis of major depressive disorder. Mol Psychiatry. doi:10.1038/s41380-023-01957-9

35. Flint, J., & Kendler, K. S. (2014). The genetics of major depression. Neuron, 81(3), 484–503. doi:10.1016/j.neuron.2014.01.027

36. Fuh, S. C., Fiori, L. M., Turecki, G., Nagy, C., & Li, Y. (2023). Multi-omic modeling of antidepressant response implicates dynamic immune and inflammatory changes in individuals who respond to treatment. PLoS One, 18(5), e0285123. doi:10.1371/journal.pone.0285123

37. Gao, C., Xu, Z., Tan, T., Chen, Z., Shen, T., Chen, L., … Yuan, Y. (2022). Combination of spontaneous regional brain activity and HTR1A/1B DNA methylation to predict early responses to antidepressant treatments in MDD. J Affect Disord, 302, 249–257. doi:10.1016/j.jad.2022.01.098

38. Garcia-Gonzalez, J., Tansey, K. E., Hauser, J., Henigsberg, N., Maier, W., Mors, O., … Fabbri, C. (2017). Pharmacogenetics of antidepressant response: A polygenic approach. Prog Neuropsychopharmacol Biol Psychiatry, 75, 128–134. doi:10.1016/j.pnpbp.2017.01.011

39. Gartlehner, G., Wagner, G., Matyas, N., Titscher, V., Greimel, J., Lux, L., … Lohr, K. N. (2017). Pharmacological and non-pharmacological treatments for major depressive disorder: review of systematic reviews. BMJ Open, 7(6), e014912. doi:10.1136/bmjopen-2016-014912

40. Gassen, N. C., Fries, G. R., Zannas, A. S., Hartmann, J., Zschocke, J., Hafner, K., … Rein, T. (2015). Chaperoning epigenetics: FKBP51 decreases the activity of DNMT1 and mediates epigenetic effects of the antidepressant paroxetine. Sci Signal, 8(404), ra119. doi:10.1126/scisignal.aac7695

41. Gasso, P., Rodriguez, N., Blazquez, A., Monteagudo, A., Boloc, D., Plana, M. T., … Mas, S. (2017). Epigenetic and genetic variants in the HTR1B gene and clinical improvement in children and adolescents treated with fluoxetine. Prog Neuropsychopharmacol Biol Psychiatry, 75, 28–34. doi:10.1016/j.pnpbp.2016.12.003

42. Giri, A. K., Bharadwaj, S., Banerjee, P., Chakraborty, S., Parekatt, V., Rajashekar, D., … Bharadwaj, D. (2017). DNA methylation profiling reveals the presence of population-specific signatures correlating with phenotypic characteristics. Mol Genet Genomics, 292(3), 655–662. doi:10.1007/s00438-017-1298-0

43. Goodwin, R. D., Dierker, L. C., Wu, M., Galea, S., Hoven, C. W., & Weinberger, A. H. (2022). Trends in U.S. Depression Prevalence From 2015 to 2020: The Widening Treatment Gap. Am J Prev Med, 63(5), 726–733. doi:10.1016/j.amepre.2022.05.014

44. Guo, J. U., Su, Y., Shin, J. H., Shin, J., Li, H., Xie, B., … Song, H. (2014). Distribution, recognition and regulation of non-CpG methylation in the adult mammalian brain. Nat Neurosci, 17(2), 215–222. doi:10.1038/nn.3607

45. Hamon, M., & Blier, P. (2013). Monoamine neurocircuitry in depression and strategies for new treatments. Prog Neuropsychopharmacol Biol Psychiatry, 45, 54–63. doi:10.1016/j.pnpbp.2013.04.009

46. Hannon, E., Lunnon, K., Schalkwyk, L., & Mill, J. (2015). Interindividual methylomic variation across blood, cortex, and cerebellum: implications for epigenetic studies of neurological and neuropsychiatric phenotypes. Epigenetics, 10(11), 1024–1032. doi:10.1080/15592294.2015.1100786

47. Hardeveld, F., Spijker, J., De Graaf, R., Hendriks, S. M., Licht, C. M., Nolen, W. A., … Beekman, A. T. (2013). Recurrence of major depressive disorder across different treatment settings: results from the NESDA study. J Affect Disord, 147(1-3), 225–231. doi:10.1016/j.jad.2012.11.008

48. Hasin, D. S., Sarvet, A. L., Meyers, J. L., Saha, T. D., Ruan, W. J., Stohl, M., & Grant, B. F. (2018). Epidemiology of Adult DSM-5 Major Depressive Disorder and Its Specifiers in the United States. JAMA Psychiatry, 75(4), 336–346. doi:10.1001/jamapsychiatry.2017.4602

49. Heim, C., & Binder, E. B. (2012). Current research trends in early life stress and depression: review of human studies on sensitive periods, gene-environment interactions, and epigenetics. Exp Neurol, 233(1), 102–111. doi:10.1016/j.expneurol.2011.10.032

50. Hou, L., Zhang, X., Wang, D., & Baccarelli, A. (2012). Environmental chemical exposures and human epigenetics. Int J Epidemiol, 41(1), 79–105. doi:10.1093/ije/dyr154

51. Hsieh, M. T., Lin, C. C., Lee, C. T., & Huang, T. L. (2019). Abnormal Brain-Derived Neurotrophic Factor Exon IX Promoter Methylation, Protein, and mRNA Levels in Patients with Major Depressive Disorder. J Clin Med, 8(5). doi:10.3390/jcm8050568

52. Humphreys, K. L., Moore, S. R., Davis, E. G., MacIsaac, J. L., Lin, D. T. S., Kobor, M. S., & Gotlib, I. H. (2019). DNA methylation of HPA-axis genes and the onset of major depressive disorder in adolescent girls: a prospective analysis. Transl Psychiatry, 9(1), 245. doi:10.1038/s41398-019-0582-7

53. Iga, J., Watanabe, S. Y., Numata, S., Umehara, H., Nishi, A., Kinoshita, M., … Ohmori, T. (2016). Association study of polymorphism in the serotonin transporter gene promoter, methylation profiles, and expression in patients with major depressive disorder. Hum Psychopharmacol, 31(3), 193–199. doi:10.1002/hup.2527

54. Jannati, A., Oberman, L. M., Rotenberg, A., & Pascual-Leone, A. (2023). Assessing the mechanisms of brain plasticity by transcranial magnetic stimulation. Neuropsychopharmacology, 48(1), 191–208. doi:10.1038/s41386-022-01453-8

55. Jia, H., Zack, M. M., Thompson, W. W., Crosby, A. E., & Gottesman, II. (2015). Impact of depression on quality-adjusted life expectancy (QALE) directly as well as indirectly through suicide. Soc Psychiatry Psychiatr Epidemiol, 50(6), 939–949. doi:10.1007/s00127-015-1019-0

56. Jin, Y., Sun, L. H., Yang, W., Cui, R. J., & Xu, S. B. (2019). The Role of BDNF in the Neuroimmune Axis Regulation of Mood Disorders. Front Neurol, 10, 515. doi:10.3389/fneur.2019.00515

57. Ju, C., Fiori, L. M., Belzeaux, R., Theroux, J. F., Chen, G. G., Aouabed, Z., … Turecki, G. (2019). Integrated genome-wide methylation and expression analyses reveal functional predictors of response to antidepressants. Transl Psychiatry, 9(1), 254. doi:10.1038/s41398-019-0589-0

58. Kang, H. J., Kim, J. M., Stewart, R., Kim, S. Y., Bae, K. Y., Kim, S. W., … Yoon, J. S. (2013). Association of SLC6A4 methylation with early adversity, characteristics and outcomes in depression. Prog Neuropsychopharmacol Biol Psychiatry, 44, 23–28. doi:10.1016/j.pnpbp.2013.01.006

59. Kendler, K. S., Gatz, M., Gardner, C. O., & Pedersen, N. L. (2006). A Swedish national twin study of lifetime major depression. Am J Psychiatry, 163(1), 109–114. doi:10.1176/appi.ajp.163.1.109

60. Kendler, K. S., Ohlsson, H., Lichtenstein, P., Sundquist, J., & Sundquist, K. (2018). The Genetic Epidemiology of Treated Major Depression in Sweden. Am J Psychiatry, 175(11), 1137–1144. doi:10.1176/appi.ajp.2018.17111251

61. Kim, I. B., Lee, J. H., & Park, S. C. (2022). The Relationship between Stress, Inflammation, and Depression. Biomedicines, 10(8). doi:10.3390/biomedicines10081929

62. Kim, J. K., Samaranayake, M., & Pradhan, S. (2009). Epigenetic mechanisms in mammals. Cell Mol Life Sci, 66(4), 596–612. doi:10.1007/s00018-008-8432-4

63. Kim, J. M., Stewart, R., Kang, H. J., Bae, K. Y., Kim, S. W., Shin, I. S., … Yoon, J. S. (2015). BDNF methylation and depressive disorder in acute coronary syndrome: The K-DEPACS and EsDEPACS studies. Psychoneuroendocrinology, 62, 159–165. doi:10.1016/j.psyneuen.2015.08.013

64. Kishi, T., Yoshimura, R., Ikuta, T., & Iwata, N. (2017). Brain-Derived Neurotrophic Factor and Major Depressive Disorder: Evidence from Meta-Analyses. Front Psychiatry, 8, 308. doi:10.3389/fpsyt.2017.00308

65. Kleimann, A., Kotsiari, A., Sperling, W., Groschl, M., Heberlein, A., Kahl, K. G., … Frieling, H. (2015). BDNF serum levels and promoter methylation of BDNF exon I, IV and VI in depressed patients receiving electroconvulsive therapy. J Neural Transm (Vienna), 122(6), 925–928. doi:10.1007/s00702-014-1336-6

66. Klengel, T., & Binder, E. B. (2015). Epigenetics of Stress-Related Psychiatric Disorders and Gene x Environment Interactions. Neuron, 86(6), 1343–1357. doi:10.1016/j.neuron.2015.05.036

67. Klengel, T., Mehta, D., Anacker, C., Rex-Haffner, M., Pruessner, J. C., Pariante, C. M., … Binder, E. B. (2013). Allele-specific FKBP5 DNA demethylation mediates gene-childhood trauma interactions. Nat Neurosci, 16(1), 33–41. doi:10.1038/nn.3275

68. Kozisek, M. E., Middlemas, D., & Bylund, D. B. (2008). Brain-derived neurotrophic factor and its receptor tropomyosin-related kinase B in the mechanism of action of antidepressant therapies. Pharmacol Ther, 117(1), 30–51. doi:10.1016/j.pharmthera.2007.07.001

69. Kringel, D., Malkusch, S., & Lotsch, J. (2021). Drugs and Epigenetic Molecular Functions. A Pharmacological Data Scientometric Analysis. Int J Mol Sci, 22(14). doi:10.3390/ijms22147250

70. Lassale, C., Batty, G. D., Baghdadli, A., Jacka, F., Sanchez-Villegas, A., Kivimaki, M., & Akbaraly, T. (2019). Healthy dietary indices and risk of depressive outcomes: a systematic review and meta-analysis of observational studies. Mol Psychiatry, 24(7), 965–986. doi:10.1038/s41380-018-0237-8

71. Lee, Y., Ragguett, R. M., Mansur, R. B., Boutilier, J. J., Rosenblat, J. D., Trevizol, A., … McIntyre, R. S. (2018). Applications of machine learning algorithms to predict therapeutic outcomes in depression: A meta-analysis and systematic review. J Affect Disord, 241, 519–532. doi:10.1016/j.jad.2018.08.073

72. Li, L., Wang, T., Chen, S., Yue, Y., Xu, Z., & Yuan, Y. (2021). DNA methylations of brain-derived neurotrophic factor exon VI are associated with major depressive disorder and antidepressant-induced remission in females. J Affect Disord, 295, 101–107. doi:10.1016/j.jad.2021.08.016

73. Li, M., Yao, X., Sun, L., Zhao, L., Xu, W., Zhao, H., … Cui, R. (2020). Effects of Electroconvulsive Therapy on Depression and Its Potential Mechanism. Front Psychol, 11, 80. doi:10.3389/fpsyg.2020.00080

74. Li, Z., Ruan, M., Chen, J., & Fang, Y. (2021). Major Depressive Disorder: Advances in Neuroscience Research and Translational Applications. Neurosci Bull, 37(6), 863–880. doi:10.1007/s12264-021-00638-3

75. Lieb, K., Dreimuller, N., Wagner, S., Schlicht, K., Falter, T., Neyazi, A., … Frieling, H. (2018). BDNF Plasma Levels and BDNF Exon IV Promoter Methylation as Predictors for Antidepressant Treatment Response. Front Psychiatry, 9, 511. doi:10.3389/fpsyt.2018.00511

76. Lotsch, J., Schneider, G., Reker, D., Parnham, M. J., Schneider, P., Geisslinger, G., & Doehring, A. (2013). Common non-epigenetic drugs as epigenetic modulators. Trends Mol Med, 19(12), 742–753. doi:10.1016/j.molmed.2013.08.006

77. Loyfer, N., Magenheim, J., Peretz, A., Cann, G., Bredno, J., Klochendler, A., … Kaplan, T. (2023). A DNA methylation atlas of normal human cell types. Nature, 613(7943), 355–364. doi:10.1038/s41586-022-05580-6

78. Luo, L., Hu, J., Huang, R., Kong, D., Hu, W., Ding, Y., & Yu, H. (2023). The association between dietary inflammation index and depression. Front Psychiatry, 14, 1131802. doi:10.3389/fpsyt.2023.1131802

79. Maier, H. B., Moschny, N., Eberle, F., Jahn, K., Folsche, T., Schulke, R., … Neyazi, A. (2023). DNA Methylation of POMC and NR3C1-1F and Its Implication in Major Depressive Disorder and Electroconvulsive Therapy. Pharmacopsychiatry, 56(2), 64–72. doi:10.1055/a-2034-6536

80. Martinez-Pinteno, A., Rodriguez, N., Blazquez, A., Plana, M. T., Varela, E., Gasso, P., … Mas, S. (2021). DNA Methylation of Fluoxetine Response in Child and Adolescence: Preliminary Results. Pharmgenomics Pers Med, 14, 459–467. doi:10.2147/PGPM.S289480

81. Martins, J., Czamara, D., Sauer, S., Rex-Haffner, M., Dittrich, K., Dorr, P., … Binder, E. B. (2021). Childhood adversity correlates with stable changes in DNA methylation trajectories in children and converges with epigenetic signatures of prenatal stress. Neurobiol Stress, 15, 100336. doi:10.1016/j.ynstr.2021.100336

82. Maugeri, A., & Barchitta, M. (2020). How Dietary Factors Affect DNA Methylation: Lesson from Epidemiological Studies. Medicina (Kaunas), 56(8). doi:10.3390/medicina56080374

83. McEwen, B. S., Nasca, C., & Gray, J. D. (2016). Stress Effects on Neuronal Structure: Hippocampus, Amygdala, and Prefrontal Cortex. Neuropsychopharmacology, 41(1), 3–23. doi:10.1038/npp.2015.171

84. McGuffin, P., Rijsdijk, F., Andrew, M., Sham, P., Katz, R., & Cardno, A. (2003). The heritability of bipolar affective disorder and the genetic relationship to unipolar depression. Arch Gen Psychiatry, 60(5), 497–502. doi:10.1001/archpsyc.60.5.497

85. Meerman, J. J., Ter Hark, S. E., Janzing, J. G. E., & Coenen, M. J. H. (2022). The Potential of Polygenic Risk Scores to Predict Antidepressant Treatment Response in Major Depression: A Systematic Review. J Affect Disord, 304, 1–11. doi:10.1016/j.jad.2022.02.015

86. Moffitt, T. E., Caspi, A., Taylor, A., Kokaua, J., Milne, B. J., Polanczyk, G., & Poulton, R. (2010). How common are common mental disorders? Evidence that lifetime prevalence rates are doubled by prospective versus retrospective ascertainment. Psychol Med, 40(6), 899–909. doi:10.1017/S0033291709991036

87. Mohammadi, S., Beh-Pajooh, A., Ahmadimanesh, M., Amini, M., Ghazi-Khansari, M., Moallem, S. A., … Ghahremani, M. H. (2022). Evaluation of DNA methylation in BDNF, SLC6A4, NR3C1 and FKBP5 before and after treatment with selective serotonin-reuptake inhibitor in major depressive disorder. Epigenomics, 14(20), 1269–1280. doi:10.2217/epi-2022-0246

88. Moon, Y. K., Kim, H., Kim, S., Lim, S. W., & Kim, D. K. (2023). Influence of antidepressant treatment on SLC6A4 methylation in Korean patients with major depression. Am J Med Genet B Neuropsychiatr Genet, 192(1-2), 28–37. doi:10.1002/ajmg.b.32921

89. Morilak, D. A., & Frazer, A. (2004). Antidepressants and brain monoaminergic systems: a dimensional approach to understanding their behavioural effects in depression and anxiety disorders. Int J Neuropsychopharmacol, 7(2), 193–218. doi:10.1017/S1461145704004080

90. Moschny, N., Jahn, K., Bajbouj, M., Maier, H. B., Ballmaier, M., Khan, A. Q., … Neyazi, A. (2020). DNA Methylation of the t-PA Gene Differs Between Various Immune Cell Subtypes Isolated From Depressed Patients Receiving Electroconvulsive Therapy. Front Psychiatry, 11, 571. doi:10.3389/fpsyt.2020.00571

91. Moschny, N., Zindler, T., Jahn, K., Dorda, M., Davenport, C. F., Wiehlmann, L., … Frieling, H. (2020). Novel candidate genes for ECT response prediction-a pilot study analyzing the DNA methylome of depressed patients receiving electroconvulsive therapy. Clin Epigenetics, 12(1), 114. doi:10.1186/s13148-020-00891-9

92. Neyazi, A., Theilmann, W., Brandt, C., Rantamaki, T., Matsui, N., Rhein, M., … Loscher, W. (2018). P11 promoter methylation predicts the antidepressant effect of electroconvulsive therapy. Transl Psychiatry, 8(1), 25. doi:10.1038/s41398-017-0077-3

93. Ng, C. H., Easteal, S., Tan, S., Schweitzer, I., Ho, B. K., & Aziz, S. (2006). Serotonin transporter polymorphisms and clinical response to sertraline across ethnicities. Prog Neuropsychopharmacol Biol Psychiatry, 30(5), 953–957. doi:10.1016/j.pnpbp.2006.02.015

94. Nishiyama, A., & Nakanishi, M. (2021). Navigating the DNA methylation landscape of cancer. Trends Genet, 37(11), 1012–1027. doi:10.1016/j.tig.2021.05.002

95. Okada, S., Morinobu, S., Fuchikami, M., Segawa, M., Yokomaku, K., Kataoka, T., … Mimura, M. (2014). The potential of SLC6A4 gene methylation analysis for the diagnosis and treatment of major depression. J Psychiatr Res, 53, 47–53. doi:10.1016/j.jpsychires.2014.02.002

96. Ottenhof, K. W., Sild, M., Levesque, M. L., Ruhe, H. G., & Booij, L. (2018). TPH2 polymorphisms across the spectrum of psychiatric morbidity: A systematic review and meta-analysis. Neurosci Biobehav Rev, 92, 29–42. doi:10.1016/j.neubiorev.2018.05.018

97. Ousdal, O. T., Brancati, G. E., Kessler, U., Erchinger, V., Dale, A. M., Abbott, C., & Oltedal, L. (2022). The Neurobiological Effects of Electroconvulsive Therapy Studied Through Magnetic Resonance: What Have We Learned, and Where Do We Go? Biol Psychiatry, 91(6), 540–549. doi:10.1016/j.biopsych.2021.05.023

98. Ownby, R. L., Crocco, E., Acevedo, A., John, V., & Loewenstein, D. (2006). Depression and risk for Alzheimer disease: systematic review, meta-analysis, and metaregression analysis. Arch Gen Psychiatry, 63(5), 530–538. doi:10.1001/archpsyc.63.5.530

99. Papakostas, G. I. (2008). Tolerability of modern antidepressants. J Clin Psychiatry, 69 *Suppl E1*, 8–13. Retrieved from https://www.ncbi.nlm.nih.gov/pubmed/18494538

100. Pariante, C. M. (2017). Why are depressed patients inflamed? A reflection on 20 years of research on depression, glucocorticoid resistance and inflammation. Eur Neuropsychopharmacol, 27(6), 554–559. doi:10.1016/j.euroneuro.2017.04.001

101. Penner-Goeke, S., & Binder, E. B. (2019). Epigenetics and depression Dialogues Clin Neurosci, 21(4), 397–405. doi:10.31887/DCNS.2019.21.4/ebinder

102. Petralia, M. C., Mazzon, E., Fagone, P., Basile, M. S., Lenzo, V., Quattropani, M. C., … Nicoletti, F. (2020). The cytokine network in the pathogenesis of major depressive disorder. Close to translation? Autoimmun Rev, 19(5), 102504. doi:10.1016/j.autrev.2020.102504

103. Powell, T. R., Smith, R. G., Hackinger, S., Schalkwyk, L. C., Uher, R., McGuffin, P., … Tansey, K. E. (2013). DNA methylation in interleukin-11 predicts clinical response to antidepressants in GENDEP. Transl Psychiatry, 3(9), e300. doi:10.1038/tp.2013.73

104. Provencal, N., Arloth, J., Cattaneo, A., Anacker, C., Cattane, N., Wiechmann, T., … Binder, E. B. (2020). Glucocorticoid exposure during hippocampal neurogenesis primes future stress response by inducing changes in DNA methylation. Proc Natl Acad Sci U S A, 117(38), 23280–23285. doi:10.1073/pnas.1820842116

105. Psych, E. C., Akbarian, S., Liu, C., Knowles, J. A., Vaccarino, F. M., Farnham, P. J., … Sestan, N. (2015). The PsychENCODE project. Nat Neurosci, 18(12), 1707–1712. doi:10.1038/nn.4156

106. Qiao, M., Jiang, Q. S., Liu, Y. J., Hu, X. Y., Wang, L. J., Zhou, Q. X., & Qiu, H. M. (2019). Antidepressant mechanisms of venlafaxine involving increasing histone acetylation and modulating tyrosine hydroxylase and tryptophan hydroxylase expression in hippocampus of depressive rats. Neuroreport, 30(4), 255–261. doi:10.1097/WNR.0000000000001191

107. Ridout, K. K., Ridout, S. J., Price, L. H., Sen, S., & Tyrka, A. R. (2016). Depression and telomere length: A meta-analysis. J Affect Disord, 191, 237–247. doi:10.1016/j.jad.2015.11.052

108. Roadmap Epigenomics, C., Kundaje, A., Meuleman, W., Ernst, J., Bilenky, M., Yen, A., … Kellis, M. (2015). Integrative analysis of 111 reference human epigenomes. Nature, 518(7539), 317–330. doi:10.1038/nature14248

109. Roberts, S., Lester, K. J., Hudson, J. L., Rapee, R. M., Creswell, C., Cooper, P. J., … Eley, T. C. (2014). Serotonin transporter [corrected] methylation and response to cognitive behaviour therapy in children with anxiety disorders. Transl Psychiatry, 4(9), e444. doi:10.1038/tp.2014.83

110. Rotella, F., & Mannucci, E. (2013). Depression as a risk factor for diabetes: a meta-analysis of longitudinal studies. J Clin Psychiatry, 74(1), 31–37. doi:10.4088/JCP.12r07922

111. Roy, B., Dunbar, M., Shelton, R. C., & Dwivedi, Y. (2017). Identification of MicroRNA-124-3p as a Putative Epigenetic Signature of Major Depressive Disorder. Neuropsychopharmacology, 42(4), 864–875. doi:10.1038/npp.2016.175

112. Rush, A. J., Fava, M., Wisniewski, S. R., Lavori, P. W., Trivedi, M. H., Sackeim, H. A., … Group, S. D. I. (2004). Sequenced treatment alternatives to relieve depression (STAR*D): rationale and design. Control Clin Trials, 25(1), 119–142. doi:10.1016/s0197-2456(03)00112-0

113. Sailani, M. R., Halling, J. F., Moller, H. D., Lee, H., Plomgaard, P., Pilegaard, H., … Regenberg, B. (2019). Lifelong physical activity is associated with promoter hypomethylation of genes involved in metabolism, myogenesis, contractile properties and oxidative stress resistance in aged human skeletal muscle. Sci Rep, 9(1), 3272. doi:10.1038/s41598-018-37895-8

114. Schiele, M. A., Kollert, L., Lesch, K. P., Arolt, V., Zwanzger, P., Deckert, J., … Domschke, K. (2019). Hypermethylation of the serotonin transporter gene promoter in panic disorder-Epigenetic imprint of comorbid depression? Eur Neuropsychopharmacol, 29(10), 1161–1167. doi:10.1016/j.euroneuro.2019.07.131

115. Schiele, M. A., Zwanzger, P., Schwarte, K., Arolt, V., Baune, B. T., & Domschke, K. (2021). Serotonin Transporter Gene Promoter Hypomethylation as a Predictor of Antidepressant Treatment Response in Major Depression: A Replication Study. Int J Neuropsychopharmacol, 24(3), 191–199. doi:10.1093/ijnp/pyaa081

116. Schildkraut, J. J. (1965). The catecholamine hypothesis of affective disorders: a review of supporting evidence. Am J Psychiatry, 122(5), 509–522. doi:10.1176/ajp.122.5.509

117. Schurgers, G., Walter, S., Pishva, E., Guloksuz, S., Peerbooms, O., Incio, L. R., … Rutten, B. P. F. (2022). Longitudinal alterations in mRNA expression of the BDNF neurotrophin signaling cascade in blood correlate with changes in depression scores in patients undergoing electroconvulsive therapy. Eur Neuropsychopharmacol, 63, 60–70. doi:10.1016/j.euroneuro.2022.07.183

118. Shadrina, M., Bondarenko, E. A., & Slominsky, P. A. (2018). Genetics Factors in Major Depression Disease. Front Psychiatry, 9, 334. doi:10.3389/fpsyt.2018.00334

119. Shen, T., Li, X., Chen, L., Chen, Z., Tan, T., Hua, T., … Xu, Z. (2020). The relationship of tryptophan hydroxylase-2 methylation to early-life stress and its impact on short-term antidepressant treatment response. J Affect Disord, 276, 850–858. doi:10.1016/j.jad.2020.07.111

120. Sirignano, L., Frank, J., Kranaster, L., Witt, S. H., Streit, F., Zillich, L., … Foo, J. C. (2021). Methylome-wide change associated with response to electroconvulsive therapy in depressed patients. Transl Psychiatry, 11(1), 347. doi:10.1038/s41398-021-01474-9

121. Souery, D., Oswald, P., Massat, I., Bailer, U., Bollen, J., Demyttenaere, K., … Group for the Study of Resistant, D. (2007). Clinical factors associated with treatment resistance in major depressive disorder: results from a European multicenter study. J Clin Psychiatry, 68(7), 1062–1070. doi:10.4088/jcp.v68n0713

122. Surtees, P. G., Wainwright, N. W., Luben, R. N., Wareham, N. J., Bingham, S. A., & Khaw, K. T. (2008). Depression and ischemic heart disease mortality: evidence from the EPIC-Norfolk United Kingdom prospective cohort study. Am J Psychiatry, 165(4), 515–523. doi:10.1176/appi.ajp.2007.07061018

123. Tadic, A., Muller-Engling, L., Schlicht, K. F., Kotsiari, A., Dreimuller, N., Kleimann, A., … Frieling, H. (2014). Methylation of the promoter of brain-derived neurotrophic factor exon IV and antidepressant response in major depression. Mol Psychiatry, 19(3), 281–283. doi:10.1038/mp.2013.58

124. Takeuchi, N., Nonen, S., Kato, M., Wakeno, M., Takekita, Y., Kinoshita, T., & Kugawa, F. (2017). Therapeutic Response to Paroxetine in Major Depressive Disorder Predicted by DNA Methylation. Neuropsychobiology, 75(2), 81–88. doi:10.1159/000480512

125. Tayyab, M., Shahi, M. H., Farheen, S., Mariyath, M. P. M., Khanam, N., Castresana, J. S., & Hossain, M. M. (2018). Sonic hedgehog, Wnt, and brain-derived neurotrophic factor cell signaling pathway crosstalk: potential therapy for depression. J Neurosci Res, 96(1), 53–62. doi:10.1002/jnr.24104

126. Thomas, L., Kessler, D., Campbell, J., Morrison, J., Peters, T. J., Williams, C., … Wiles, N. (2013). Prevalence of treatment-resistant depression in primary care: cross-sectional data. Br J Gen Pract, 63(617), e852–858. doi:10.3399/bjgp13X675430

127. Tiger, M., Varnas, K., Okubo, Y., & Lundberg, J. (2018). The 5-HT(1B) receptor – a potential target for antidepressant treatment. Psychopharmacology (Berl), 235(5), 1317–1334. doi:10.1007/s00213-018-4872-1

128. Tong, J., Meyer, J. H., Furukawa, Y., Boileau, I., Chang, L. J., Wilson, A. A., … Kish, S. J. (2013). Distribution of monoamine oxidase proteins in human brain: implications for brain imaging studies. J Cereb Blood Flow Metab, 33(6), 863–871. doi:10.1038/jcbfm.2013.19

129. Trivedi, M. H., Rush, A. J., Wisniewski, S. R., Nierenberg, A. A., Warden, D., Ritz, L., … Team, S. D. S. (2006). Evaluation of outcomes with citalopram for depression using measurement-based care in STAR*D: implications for clinical practice. Am J Psychiatry, 163(1), 28–40. doi:10.1176/appi.ajp.163.1.28

130. Tsai, P. C., & Bell, J. T. (2015). Power and sample size estimation for epigenome-wide association scans to detect differential DNA methylation. Int J Epidemiol, 44(4), 1429–1441. doi:10.1093/ije/dyv041

131. Tsai, S. J., Hong, C. J., Liou, Y. J., Yu, Y. W., Chen, T. J., Hou, S. J., & Yen, F. C. (2009). Tryptophan hydroxylase 2 gene is associated with major depression and antidepressant treatment response. Prog Neuropsychopharmacol Biol Psychiatry, 33(4), 637–641. doi:10.1016/j.pnpbp.2009.02.020

132. Undurraga, J., & Baldessarini, R. J. (2012). Randomized, placebo-controlled trials of antidepressants for acute major depression: thirty-year meta-analytic review. Neuropsychopharmacology, 37(4), 851–864. doi:10.1038/npp.2011.306

133. van den Oord, C., Copeland, W. E., Zhao, M., Xie, L. Y., Aberg, K. A., & van den Oord, E. (2022). DNA methylation signatures of childhood trauma predict psychiatric disorders and other adverse outcomes 17 years after exposure. Mol Psychiatry, 27(8), 3367–3373. doi:10.1038/s41380-022-01597-5

134. van der Knaap, L. J., Riese, H., Hudziak, J. J., Verbiest, M. M., Verhulst, F. C., Oldehinkel, A. J., & van Oort, F. V. (2014). Glucocorticoid receptor gene (NR3C1) methylation following stressful events between birth and adolescence. The TRAILS study. Transl Psychiatry, 4(4), e381. doi:10.1038/tp.2014.22

135. Vancampfort, D., Mitchell, A. J., De Hert, M., Sienaert, P., Probst, M., Buys, R., & Stubbs, B. (2015). Type 2 Diabetes in Patients with Major Depressive Disorder: A Meta-Analysis of Prevalence Estimates and Predictors. Depress Anxiety, 32(10), 763–773. doi:10.1002/da.22387

136. Walther, D. J., Peter, J. U., Bashammakh, S., Hortnagl, H., Voits, M., Fink, H., & Bader, M. (2003). Synthesis of serotonin by a second tryptophan hydroxylase isoform. Science, 299(5603), 76. doi:10.1126/science.1078197

137. Wang, P., Lv, Q., Mao, Y., Zhang, C., Bao, C., Sun, H., … Fang, Y. (2018). HTR1A/1B DNA methylation may predict escitalopram treatment response in depressed Chinese Han patients. J Affect Disord, 228, 222–228. doi:10.1016/j.jad.2017.12.010

138. Wang, P., Zhang, C., Lv, Q., Bao, C., Sun, H., Ma, G., … Cai, W. (2018). Association of DNA methylation in BDNF with escitalopram treatment response in depressed Chinese Han patients. Eur J Clin Pharmacol, 74(8), 1011–1020. doi:10.1007/s00228-018-2463-z

139. Warden, D., Rush, A. J., Trivedi, M. H., Fava, M., & Wisniewski, S. R. (2007). The STAR*D Project results: a comprehensive review of findings. Curr Psychiatry Rep, 9(6), 449–459. doi:10.1007/s11920-007-0061-3

140. Weaver, I. C., Meaney, M. J., & Szyf, M. (2006). Maternal care effects on the hippocampal transcriptome and anxiety-mediated behaviors in the offspring that are reversible in adulthood. Proc Natl Acad Sci U S A, 103(9), 3480–3485. doi:10.1073/pnas.0507526103

141. Wittenberg, G. M., Stylianou, A., Zhang, Y., Sun, Y., Gupta, A., Jagannatha, P. S., … Drevets, W. C. (2020). Effects of immunomodulatory drugs on depressive symptoms: A mega-analysis of randomized, placebo-controlled clinical trials in inflammatory disorders. Mol Psychiatry, 25(6), 1275–1285. doi:10.1038/s41380-019-0471-8

142. Wong, C. C., Caspi, A., Williams, B., Craig, I. W., Houts, R., Ambler, A., … Mill, J. (2010). A longitudinal study of epigenetic variation in twins. Epigenetics, 5(6), 516–526. doi:10.4161/epi.5.6.12226

143. Xu, Z., Chen, Z., Shen, T., Chen, L., Tan, T., Gao, C., … Zhang, Z. (2022). The impact of HTR1A and HTR1B methylation combined with stress/genotype on early antidepressant efficacy. Psychiatry Clin Neurosci, 76(2), 51–57. doi:10.1111/pcn.13314

144. Yeung, A. W. K., Georgieva, M. G., Atanasov, A. G., & Tzvetkov, N. T. (2019). Monoamine Oxidases (MAOs) as Privileged Molecular Targets in Neuroscience: Research Literature Analysis. Front Mol Neurosci, 12, 143. doi:10.3389/fnmol.2019.00143

145. Zhang, Y., Chang, Z., Chen, J., Ling, Y., Liu, X., Feng, Z., … Zhang, C. (2015). Methylation of the tryptophan hydroxylase–2 gene is associated with mRNA expression in patients with major depression with suicide attempts. Mol Med Rep, 12(2), 3184–3190. doi:10.3892/mmr.2015.3748

